# Scalable Bayesian functional GWAS method accounting for multivariate quantitative functional annotations with applications to studying Alzheimer’s disease

**DOI:** 10.1101/2022.08.12.22278704

**Authors:** Junyu Chen, Lei Wang, Philip L. De Jager, David A. Bennett, Aron S. Buchman, Jingjing Yang

## Abstract

Existing methods for integrating functional annotations in GWAS to fine-map and prioritize potential causal variants are either limited to using non-overlapped categorical annotations, or limited by the computation burden of modeling genome-wide variants. To overcome these limitations, we propose a scalable Bayesian functional GWAS method to account for multivariate quantitative functional annotations (BFGWAS_QUANT), accompanied by a scalable computation algorithm enabling joint modeling of genome-wide variants. Simulation studies validated the performance of BFGWAS_QUANT for accurately quantifying annotation enrichment and improving GWAS power. Applying BFGWAS_QUANT to study five Alzheimer’s disease (AD) related phenotypes using individual-level GWAS data (n=∼1K), we found that histone modification annotations have higher enrichment than eQTL annotations for all considered phenotypes, with the highest enrichment in H3K27me3 (polycomb regression). We also found that cis-eQTL in microglia had higher enrichment than eQTL of bulk brain frontal cortex tissue for all considered phenotypes. A similar enrichment pattern was also identified using the IGAP summary-level GWAS data of AD (n=∼54K). The strongest known *APOE E4* risk allele was identified for all five phenotypes and the *APOE* locus was validated using the IGAP data. BFGWAS_QUANT fine-mapped 32 significant variants from 1073 genome-wide significant variants in the IGAP data. We further demonstrated that the polygenic risk scores (PRS) using effect size estimates by BFGWAS_QUANT had similar prediction accuracy as other methods assuming a sparse causal model. Overall, BFGWAS_QUANT provides a useful GWAS tool for quantifying annotation enrichment and prioritizing potential causal variants.

## Introduction

Although thousands of significant associations have been identified by single variant genome-wide association studies (GWAS) for complex traits and diseases, majority GWAS signals reside in the noncoding genome regions and have unknown biological meaning^1-3^. Existing GWAS results based on single variant tests are still difficult to interpret with respect to the underlying biological mechanisms^4; 5^. Promisingly, advancements in sequencing technology have made plenteous multi-omics data available which provide functional annotations of genetic variants available to the scientific community –– Combined Annotation-Dependent Depletion (CADD) score^6^, Roadmap Epigenomics Mapping Consortium^7^ providing DNA methylation, histone modification, and chromatin accessibility information of various human tissues, the Encyclopedia of DNA Elements (ENCODE)^8^ providing functional information of human and mouse genomes, and the Genotype-Tissue Expression (GTEx) project^9^ providing expression quantitative trait loci (eQTL) information of 54 human tissues. In particular, molecular quantitative trait loci (QTL) mapped from profiles of molecular phenotypes (e.g., gene expression from GTEx^9^, chromatin marks from Roadmap^7; 10; 11^, and protein abundances^12^) and corresponding genomes (genotype data) have been shown to be enriched with GWAS signals and help interpretate the underlying biology for studying complex traits and diseases^13-15^. Especially, these molecular QTL have been leveraged to prioritize GWAS associations of complex traits and diseases^16-20^.

An intuitive but widely used ad-hoc approach is to fine-map and prioritize potential causal GWAS signals that are also molecular QTL^21^, or located in a region with histone modifications. Recently, advanced statistical methods have been proposed to integrate non-overlapped categorical functional annotation (assigning one function label per variant) with GWAS data to fine-map GWAS results^22-26^. Additionally, PAINTOR was proposed to integrate multivariate quantitative functional annotations with GWAS summary statistics to fine-map GWAS loci with thousands of variants^27; 28^; FunSPU^29^ and STAAR^30^ were proposed to incorporate multiple biological annotations for rare variant association tests. These existing methods have shown the feasibility and promising results of integrating multivariate quantitative functional annotations with GWAS data to fine-map GWAS results and prioritize potential causal variants. However, these methods were not developed for joint modeling millions of genome-wide variants with multivariate quantitative annotations, which would lead to less accurate quantification of annotation enrichment and reduced power of fine-mapping.

Here, we propose a scalable **B**ayesian **F**unctional **GWAS** method for integrating multivariate **QUANT**itative functional annotations with GWAS data by a Bayesian hierarchical variable selection regression model, referred to as **BFGWAS_QUANT**. BFGWAS_QUANT assumes a hierarchical logistic prior for the causal probabilities of genetic variants in the standard Bayesian Variable Selection Regression (BVSR) based GWAS method^31^ to jointly model millions of genome-wide genetic variants. BFGWAS_QUANT adapts the scalable Expectation Maximization Markov Chain Monte Carlo (EM-MCMC) algorithm developed by the previous Bayesian functional GWAS (BFGWAS) method that only models non-overlapped categorical annotations (referred to as BFGWAS_CAT in this paper)^22^. BFGWAS_QUANT further improves the computation efficiency by pre-calculating linkage disequilibrium (LD) correlation matrix and single variant test Zscore statistics that are used in the MCMC algorithm, or using reference LD correlation matrix and summary-level GWAS data. Bayesian Causal Posterior Probability (CPP) and genetic effect size estimates will be generated by BFGWAS_QUANT, along with enrichment quantification of considered multivariate quantitative functional annotations. Furthermore, Bayesian estimates of genetic effect sizes can be used to derive polygenic risk scores (PRS) that account for functional annotations.

By simulation studies, we showed that our Bayesian estimates of functional enrichment converged and GWAS power was improved over the standard BVSR method without accounting for functional annotations^31^. We then applied BFGWAS_QUANT to real GWAS data for studying Alzheimer’s disease (AD) related phenotypes^32; 33^, accounting for multivariate quantitative annotations with respect to Roadmap histone modifications^7^ of brain mid-frontal gyrus and eQTL of brain frontal cortex tissue^32; 34^ and microglia cell type^35; 36^. We showed that BFGWAS_QUANT identified interesting enrichment pattern and generated fine-mapped GWAS results using both individual-level and summary-level GWAS data. We also showed that, PRS derived from BFGWAS_QUANT effect size estimates led to similar accurate AD risk prediction as other PRS methods assuming a sparse causal model.

In the following Material and Methods section, we describe an overview of the BFGWAS_QUANT method, Roadmap histone modification and eQTL based quantitative functional annotations, simulation study design, and application studies of AD. In the Results section, we describe the results of simulation and application studies. We then end with a Discussion about the advantages and limitations of BFGWAS_QUANT.

## Material and Methods

### Bayesian hierarchical variable selection regression model

BFGWAS_QUANT assumes a Bayesian hierarchical variable selection regression (BVSR)^31^ model for genome-wide variants,

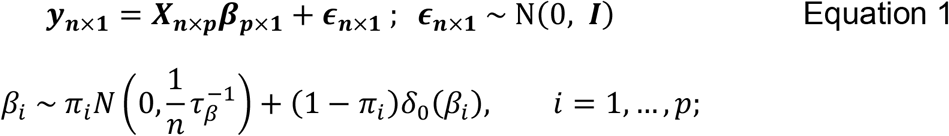

where ***y***_***n*×1**_ is a vector of standardized phenotype with *n* subjects; ***X***_***n*×*p***_ is the standardized genotype matrix with *p* genome-wide genetic variants and ***β***_***p*×1**_ is the vector of the genetic effect sizes. Spike-and-slab variable selection prior is assumed per effect size *β*_*i*_. That is, *β*_*i*_ has probability *π*_*i*_ to be non-zero and follows a normal distribution centered at zero and probability (1 − *π*_*i*_) to be zero with a point-mass density function at 0, where *π*_*i*_ denotes the “casual” probability of the *i*th variant.

We assume a hierarchical logistic model for the causal probabilities of genetic variants to account for multivariate quantitative functional annotations,

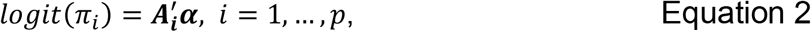

where *π*_*i*_ denotes the casual probability of the *i*th variant as in Equation 1, ***A***_*i*_ = (1, *A*_*i*1_, …, *A*_*iJ*_)′ denotes the augmented annotation vector for the *i*th variant with an intercept term as the first element, and coefficient vector ***α*** = (*α*_0_, *α*_1_, …, *α*_*J*_)′ denotes the intercept term *α*_0_ and enrichment quantification with respect to functional annotation *j* = 1, …, *J*.

Further, we assume a fixed value in the domain of (0, 1] for *τ*_*β*_, a fixed value for *α*_0_ in the domain of (−13.8, -9), and a standard normal prior for enrichment parameters (*α*_*j*_*∼N*(0, 1); *j* = 0, 1, … *J*). In particular, *τ*_*β*_ = 1 would assume the prior variance of effect sizes is the same as the marginal effect size estimates in a single variant regression model, and smaller *τ*_*β*_ values would inflate the magnitude of Bayesian effect size estimates. The lower bound value of *α*_0_ = −13.8 would assume the prior causal probability is 10^−6^ when *α*_*j*_ = 0, *j* = 1, …, *J* (see Supplementary Text for model details).

### Adapted EM-MCMC algorithm

To overcome heavy computational burden and poor mixing rate of posterior samplings by the standard MCMC algorithm^31^, we adapt the scalable EM-MCMC algorithm developed for the original BFGWAS method^22^. Specifically, we first segment genome-wide variants into approximately independent genome blocks with about 5,000∼10,000 variants based on LD structure^22; 37^. Second, conditioning on given hyper parameters (*α*_*j*_, *j* = 1, …, *J*), we conduct standard MCMC algorithm within each genome block to obtain Bayesian posterior estimates for genetic effect size (*β*_*i*_) and causal probability (*π*_*i*_) per variant (Expectation (E) step). Third, conditioning on the Bayesian estimates of (*β*_*i*_, *π*_*i*_, *i* = 1, …, *p*), we will update the values of hyper parameters (*α*_*j*_, *i* = 1, …, *J*) by maximizing their conditional posterior likelihood (Maximization (M) step). Computational optimization algorithm Broyden-Fletcher-Goldfarb-Shanno (BFGS)^38^ is used to obtain the maximum a posteriori (MAP) estimates for *α*_*j*_, *j* = 1, …, *J*. The EM steps will be iterated (∼5 iterations) until the estimates of hyper parameters converge.

The Bayesian estimate for the causal probability (*π*_*i*_) per variant is referred as CPP, and the Bayesian estimates for annotation coefficients (*α*_*j*_, *j* = 1, …, *J*) are referred as quantified enrichment of multivariate functional annotations. SNPs with CPP > 0.1068 will be considered as significantly associated with the phenotype of interest, where the significance threshold was shown equivalent to p-value < 5 **×** 10^−8^ by previously BFGWAS_CAT paper^22^. Because a multivariate regression model is fitted per genome-block, the LD among all variants per genome-block is accounted for during the Bayesian inference of CPP and effect size *β*. The GWAS results obtained by BFGWAS_QUANT will be fine-mapped and variants with enriched annotations will be prioritized.

In particular, implementing the MCMC algorithm per genome-block can greatly reduce the search space (from genome-wide to a genome-block) and facilitate parallel computing (one core per genome-block), thus leading to efficient convergence rate and improved mixing rate. Computation efficiency is further improved by implementing the MCMC algorithm using pre-calculated LD correlation matrix per genome-block and single variant test Zscore statistics, or reference LD correlation matrix and GWAS Z-score statistics, which will save up to 90% computation time compared to using individual-level GWAS data^37^. With 32 computation cores in one node, BFGWAS_QUANT can complete analyzing ∼10M SNPs in ∼4hours for 5 EM iterations.

### eQTL based functional annotations

In this paper, we considered 5 real eQTL based quantitative functional annotations. Three of these annotations (*Allcis-eQTL, 95%CredibleSet, MaxCPP*) were constructed based on standard cis-eQTL data of brain frontal cortex tissue from GTEx data^9; 34^: i) Binary annotation *Allcis-eQTL* was constructed by taking all SNPs that were identified as a significant cis-eQTL (false discovery rate (FDR) <5%; 1MB from transcription start site (TSS)) for at least one expression quantitative trait (one gene) as 1 and otherwise as 0; ii) For each gene expression trait that has at least one significant (FDR<5%) cis-eQTL, CAVIAR^16^ was used to calculate the causal posterior probability (cis-CPP) of each cis-SNP and identify a 95% credible set. SNPs that do not belong to any 95% Credible Set were taken as 0 or otherwise 1 for the annotation of *95%CredibleSet*. iii) Maximum cis-CPP per SNP across all genes were taken as quantitative values of *MaxCPP*. In addition, we took the maximum Bayesian Genome-Wide CPP of being either cis- or trans-eQTL across all genes in brain frontal cortex tissue from the Religious Orders Study and Rush Memory and Aging Project (ROS/MAP)^21; 33^ as the fourth annotation *BGW_MaxCPP*, where cis- and trans-CPPs were estimated by Bayesian genome-wide TWAS (BGW-TWAS) method^32; 37^. Lastly, we derived a fifth *Microglia-eQTL* annotation from two datasets of recent Microglia cell-type specific eQTL summary statistics^35; 39^, where 1 indicates being identified as a cis-eQTL to any gene in either Microglia dataset or otherwise 0.

### Histone modifications based functional annotation

We constructed 5 histone modification based functional annotations using the epigenomics data of core histone modifications in the brain mid frontal gyrus region from the ROADMAP Epigenomics database^7^ –– H3K4me1 (primed enhancers), H3K4me3 (promoters), H3K36me3 (gene bodies), H3K27me3 (polycomb regression) and H3K9me3 (heterochromatin). For each histone modification, peak regions from replicates of the same sample were first merged together and then overlapped with peak regions of other samples by bedtools (v 2.27.0)^40^. If a genetic variant resides in the overlapped peak regions of a histone modification, 1 would be assigned to the function annotation of such modification for this variant or 0 would be assigned otherwise.

### Simulation study design

We conducted simulation studies to validate the performance of BFGWAS_QUANT. Continuous phenotypes were simulated using the real whole genome sequence (WGS) data of Chromosomes 19 (122,745 SNPs with minor allele frequency (MAF) >0.01) for n=1,893 samples from the ROS/MAP cohort^33; 41^ and Mount Sinai Brain Bank (MSBB) study^42^. Phenotypes were simulated based on the multivariate linear additive model (Equation 1) with true genetic effect sizes ***β***_***p*×1**_ generated based on the hierarchical logistic model with multivariate quantitative functional annotations (Equation 2). Scenarios with various numbers of true causals and heritability were considered.

Besides real cis-eQTL based functional annotations of *Allcis-QTL, 95%CredibleSet, MaxCPP*, we also considered a fourth artificial annotation randomly generated from *N*(0, 1) as a negative control. With chosen annotation enrichment parameters (*α*_0_ = (−10.5, −9.5), *α*_1_ = 4, *α*_2_ = 1.5, *α*_3_ = 0.5, *α*_4_ = 0), we first calculated casual probabilities (*π*_*i*_) for all considered p=122,745 SNPs by Equation 2, where *α*_0_ was chosen to ensure the total number of true causal SNPs fall in (5, 10) with *α*_0_ = −10.5 or (15, 30) with *α*_0_ = −9.5. Second, a vector of binary indicator (*γ*_*i*_) of true causal SNPs was generated from the corresponding Bernoulli distribution with probability (*π*_*i*_) for *i* = 1, …, *p*. Third, genetic effect sizes were taken as 0 for SNPs with *γ*_*i*_ = 0 and generated from a normal distribution for SNPs with *γ*_*i*_ = 1. Finally, phenotypes were generated from Equation 1 with simulated genetic effect sizes and random errors ***ϵ****∼N*(0, (1 − *h*^2^)***I***), to ensure a target total heritability *h*^2^ = (0.25, 0.5) equally explained by all true causal SNPs. A total of four scenarios were considered, including one with relatively sparse true causals in the range of (5, 10) and one with the number of true causals in the range of (15, 30), with respect to two different heritability values (0.25, 0.5).

In addition, we considered a null enrichment scenario where none of the annotations were enriched. In this scenario, we randomly selected 10 true causal SNPs and assigned them genetic effect sizes generated from a normal distribution. We simulated the phenotype as described above with a targeted h^2^=0.5.

We repeated 100 simulations per scenario to evaluate our Bayesian estimates for annotation enrichment, total heritability, and true causal SNPs with respect to sensitivity (power) and positive predictive values (PPV)^43^. Sensitivity (power) is defined as the proportion of true positive findings among all true casual variants, and PPV is defined as the proportion of true positive findings among all identified significant associations. We took simulated true casual SNPs and those having *R*^2^ > 0.3 with true casual SNPs with 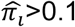 as true positive findings, following the significance rule used by the BFGWAS_CAT method^22^. The sensitivity and PPV are given by

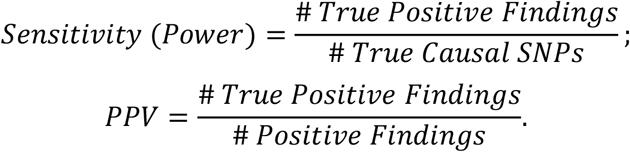

We compared with the standard BVSR method^44^ that does not account for functional annotations. We estimated the total heritability by the squared correlation between the simulated phenotypes and the PRSs based on Bayesian estimates of genetic effect sizes 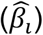 of SNPs with 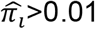,

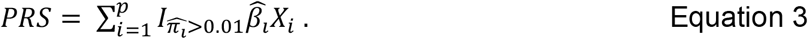

### Applications to ROS/MAP individual-level GWAS data

ROS/MAP are two prospective longitudinal community-based cohort studies, which recruit older adults without known dementia at baseline and follow participants up annually until the time of death^33; 41; 45^. Participants all agreed to annual clinical evaluation and brain autopsy at the time of death, signing an informed consent and Anatomic Gift Act. All participants in this study also signed a repository consent to allow their data to be re-purposed. WGS data were profiled for 1200 samples by using the KAPA Hyper Library Preparation Kit and Illumina HiSeq X sequencer.

We applied BFGWAS_QUANT to account for five eQTL based and five histone modifications based quantitative annotations as described above to study five AD related phenotypes^9; 32-34^, including the binary clinical diagnosis of late-onsite Alzheimer’s dementia (n=1087), three quantified postmortem pathology indices of AD (i.e., PHFtau tangle density with n=1105, β-amyloid load with n=1113, and a global measurement of AD pathology burden with n=1123), and a quantitative measurement of cognition decline rate with n=1049^33; 41^. The cognition decline rate was constructed as the random slope per sample from a linear mixed model of annual longitudinal measurements of cognition function. Details about ROS/MAP and phenotypic traits were measured can be found in previously published paper^46^. We also adjusted for the covariates of age, sex, smoking status, study index (ROS or MAP), and first 3 genotype principal components, by regressing these covariates out from the phenotypes and taking the corresponding regression residuals as the outcome in the BFGWAS_QUANT method.

### Applications to IGAP summary-level GWAS data

We applied BFGWAS_QUANT to the stage 1 summary-level GWAS data of the International Genomics of Alzheimer’s Project (IGAP)^47^, along with above 10 eQTL and histone modifications based functional annotations and reference LD generated from ROS/MAP. The stage 1 IGAP summary-level GWAS data were generated by meta-analyses consisting of 17,008 AD cases and 37,154 controls of European ancestry (n=∼54K).

### AD risk prediction by PRS

To show the usefulness of PRS for risk prediction, we evaluated two sets of PRS that were respectively derived from BFGWAS_QUANT and BVSR summary statistics using ROS/MAP GWAS data. We used the independent test samples from Mayo Clinic Alzheimer’s Disease Genetics Studies (MCADGS)^48; 49^. MCADGS contain 2,099 European-decent samples (844 AD cases and 1,255 controls) with microarray genotype data profiled that were further imputed to the 1000 Genome Project Phase^50^.

We compared PRS using Bayesian effect size estimates with three commonly used PRS methods, the standard method using informed LD-Pruning and P-value thresholding (P+T)^51; 52^, LDpred2^53^, and PRS-CS^54^. P-value thresholds of (10^−2^, 10^−3^, 10^−5^, 5 **×** 10^−8^, 10^−8^) and LD thresholds of (0.1, 0.3, 0.5, 0.7,0.9) were considered by the P+T method. Reference LD derived from 1000 Genome data were used by LDpred and PRS-CS methods. AD risk prediction accuracy was evaluated using the area under the Receiver Operating Characteristic (ROC)^55^ curve (AUC) for MCADGC test samples.

### Ethics statement

ROS/MAP and MCADGS data analyzed in this study were generated under the improvement by the Institutional Review Board (IRB) of Rush University Medical Center, Chicago, IL and Mayo Clinic, respectively. All samples analyzed in this study were de-identified and all analyses were approved by the IRB of Emory University School of Medicine.

## Results

### Simulation results

For all considered simulation scenarios, our Bayesian estimates of functional annotation enrichment achieved convergence with 4 EM iterations, as shown in boxplots of 100 simulation replicates (Figure 1 (A, D); and Supplemental Figure 1). Although BFGWAS_QUANT overestimated *α*_1_ and *α*_2_, the Bayesian estimates still reflect the correct enrichment pattern among all considered annotations. By taking the 2.5^th^ and 97.5^th^ quantiles of these 100 Bayesian estimates to estimate the corresponding 2.5^th^ and 97.5^th^ quantiles of the estimator distributions, the true enrichment values (*α*_1_ = 4, *α*_2_ = 1.5, *α*_3_ = 0.5, *α*_4_ = 0) indeed fell within this range. For example, in the scenario with *α*_0_ = −10.5 and *h*^2^ = 0.25 (Figure 1A), the estimated 2.5^th^ and 97.5^th^ quantiles are (1.^6^2, 5.50) for *α*_1_, (0.98, 4.57) for *α*_2_, (0.02, 1.20) for *α*_3_, and (0.00, 0.38) for *α*_4_. We observed precise estimates of 0 enrichment for the artificial annotation (Figure 1; Supplemental Figure 1) and all annotations in the scenario with null enrichment (Supplement Figure 3A), which demonstrated the ability of BFGWAS_QUANT to identify null enrichment.

**Figure 1.**
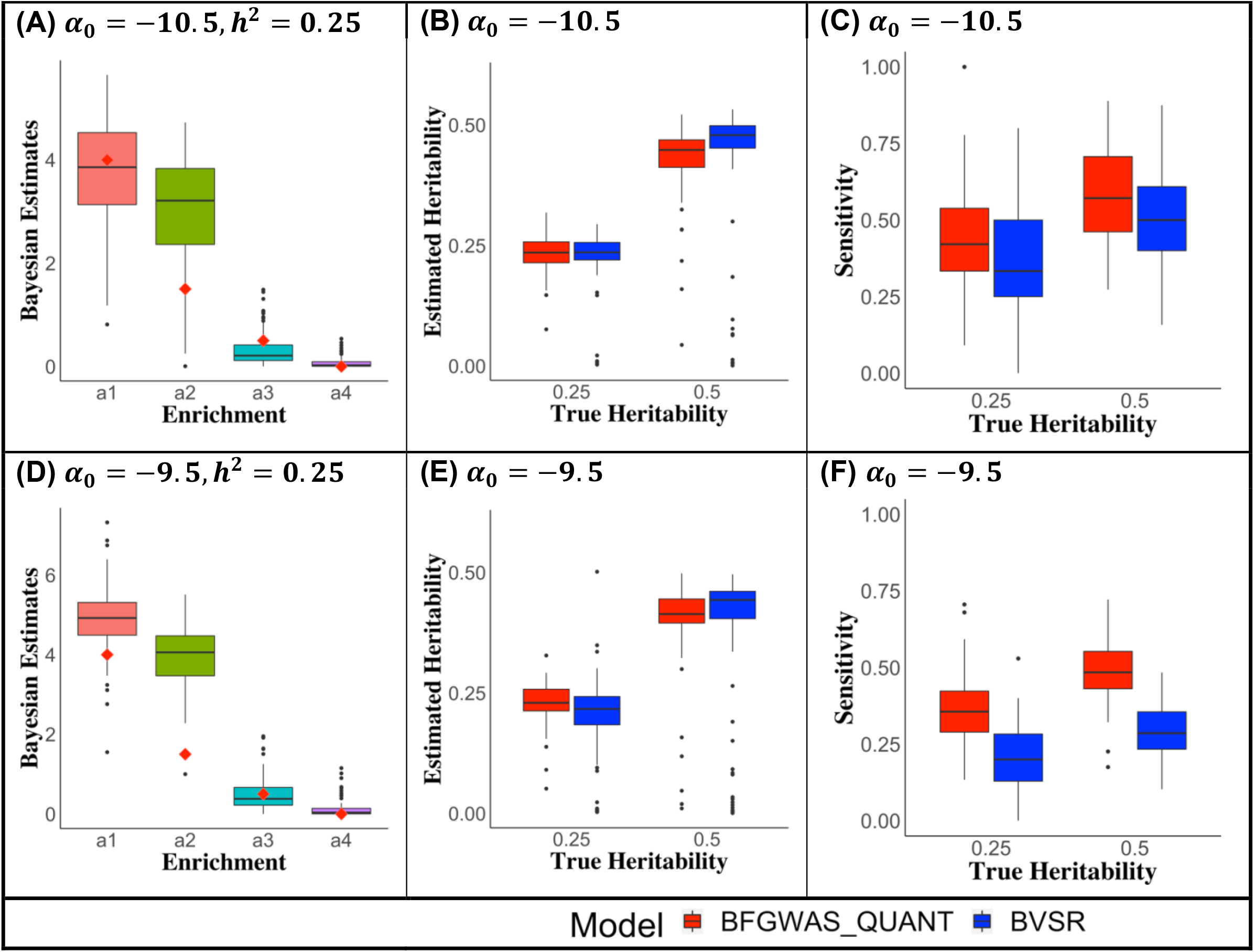
Bayesian enrichment estimates, heritability estimates, and sensitivities of simulation studies. Panels (A, B, C) are for simulations with *α*_0_ = −10.5 and panels (D, E, F) are for simulations with *α*_0_ = −9.5. Bayesian estimates of annotation enrichment (*α*_1_, *α*_2_, *α*_3_, *α*_4_) out of 100 simulations with true heritability *h*^2^ = 0.25 are shown in the respective box plot (panels A, D), where red dots denote true enrichment values. Comparable heritability estimates (panels B, E) and higher sensitivities (panels C, F) were obtained by BFGWAS_QUANT (red) vs. BVSR (blue).

By taking PRS as estimated phenotypes and taking the squared correlation between RPS and simulated phenotypes as the estimate of phenotype heritability, we obtained similar heritability estimates by BFGWAS_QUANT and BVSR, which are close to the true heritability (Figure 1 (B, E); Supplemental Figure 3B). For scenarios with true enrichment, BFGWAS_QUANT obtained substantially higher sensitivity (power) while similar positive predictive values (PPV) compared to BVSR for all scenarios (Figure 1 (C, F); Supplemental Figure 2; Supplementary Figures 4-5). For the scenario with null enrichment, both BFGWAS_QUANT and BVSR performed comparably (Supplementary Figure 3 (B, C, D)).

These simulation studies validated BFGWAS_QUANT for quantifying multivariate functional annotations, estimating phenotype heritability, and identifying true causal SNPs. By accounting for multivariate quantitative annotations, BFGWAS_QUANT showed improved performance than the standard BVSR method, especially with higher power for identifying true causal SNPs and accurate enrichment estimation.

### Application GWAS results for studying AD

Applying BFGWAS_QUANT to the individual-level ROS/MAP and summary-level IGAP GWAS data, we obtained consistent patterns for Bayesian enrichment estimates of 5 eQTL based and 5 histone modifications based functional annotations (Figure 2; Supplement Figures 6-7). In particular, the histone modifications based functional annotations had higher enrichment than eQTL based annotations when studying the individual-level ROS/MAP GWAS data, with the highest enrichment for *H3K27me3* and second highest for *H3K4me1*. Interestingly, BFGWAS_QUANT estimated the second highest enrichment for the *Microglia_eQTL* annotation when the IGAP GWAS data with a large sample size (n=∼54K). Even with a small sample size in the individual-level ROS/MAP GWAS data, the *Microglia_eQTL* annotation was still identified with higher enrichment than other annotations based on eQTL of the bulk brain frontal cortex tissue. There is mounting evidence showing that Microglia (composing <10% cells in the bulk brain frontal cortex tissues) plays important roles in the development and progression of AD pathology^56^, and cell-type specific differential expression of GWAS risk genes of AD are only present in Microglia^57^.

**Figure 2.**
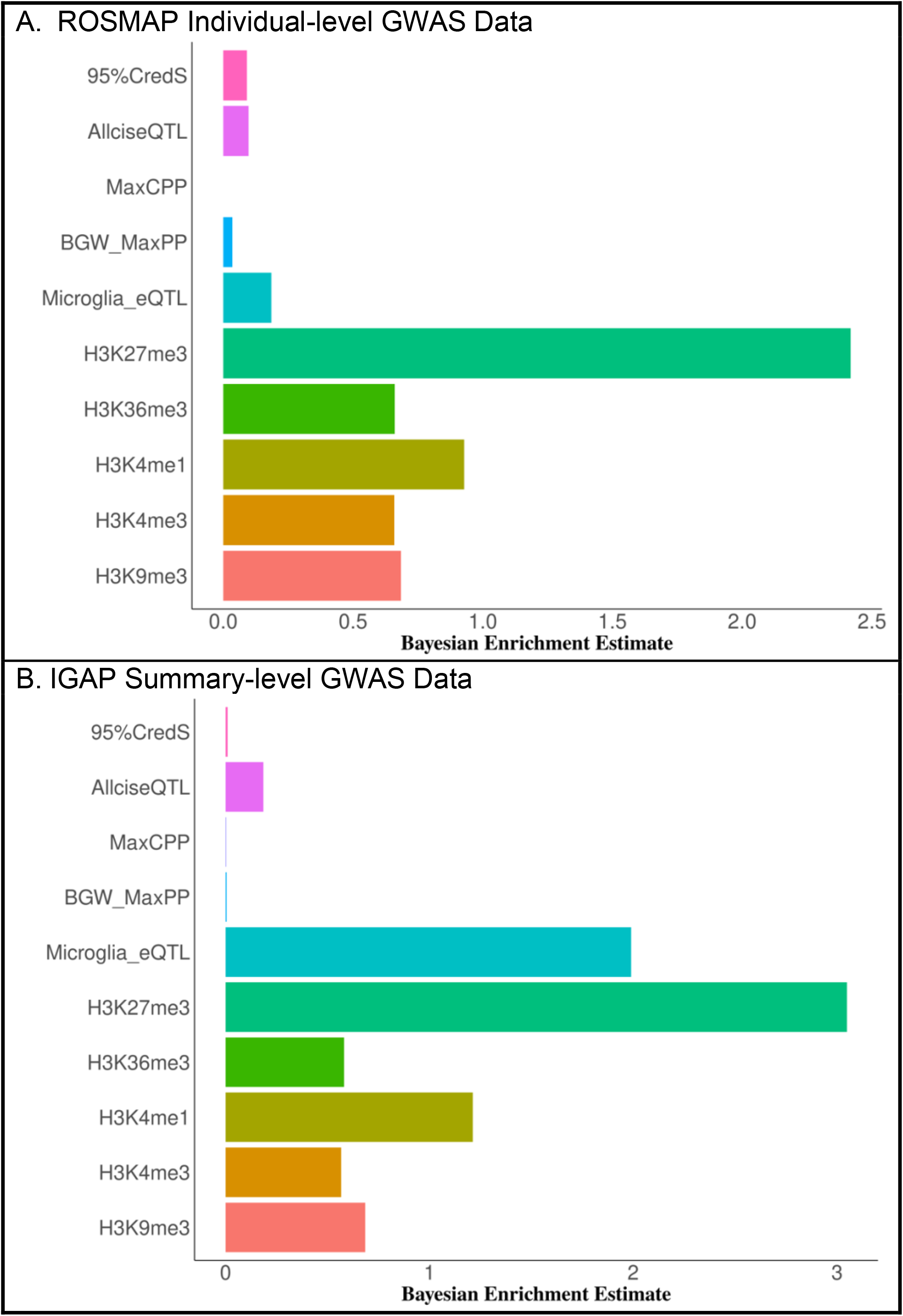
Bayesian estimates of functional annotation enrichment for Alzheimer’s dementia by using ROS/MAP individual-level (A) and IGAP summary-level (B) GWAS data. Histone modification H3K27me3 (polycomb regression) and Microglia cis-eQTL annotations were found to be most enriched for association signals of AD.

Using the ROSMAP individual-level GWAS data, four significant SNPs with Bayesian CPP > 0.1068 were identified for AD (*rs429358, rs10414043, rs769449* and *rs7256200*) by BFGWAS_QUANT (Table 1; Figure 3A; Supplemental Figures 8-9). In particular, SNP *rs429358* (CPP = 0.144, p-value = 7.72 × 10^−13^, missense variant) is the famous known *APOE E4* risk allele of AD^58^ and have significant Bayesian CPP > 0.1068 for all 5 AD related phenotypes. SNPs *rs10414043* (CPP = 0.111, p-value = 2.71 × 10^−12^) and *rs7256200* (CPP = 0.315, p-value = 2.71 × 10^−12^, regulatory variant) are upstream of the known risk gene *APOC1* of AD and blood protein traits^37; 59; 60^. Besides the missense *APOE E4* risk alleles *rs429358* and *rs7412*, one additional significant SNP *rs1065853* is identified upstream of *APOC1* for global AD pathology, which is a known GWAS signal for blood protein traits such as low-density lipoprotein^60; 61^. Out of 11 significant SNPs identified for at least one AD related phenotype (Table 1), 2 of which are intergenic and the other 9 SNPs are intron, regulatory, missense, and stop gained variants.

**Table 1.** Significant SNPs with Bayesian CPP > 0. 1068 by BFGWAS_QUANT for studying AD related phenotypes using the ROS/MAP individual-level GWAS data. SNPs with single variant test P-value <5 **×** 10^−8^ were shaded in gray.

| CHR | rsID | Gene | Function | MAF | CPP | Beta | P-value | Phenotype |
| --- | --- | --- | --- | --- | --- | --- | --- | --- |
| 1 | rs148348738 | SPATA6 | Intron | 0.011 | 0.149 | -0.039 | 4.47E-07 | Cognition decline rate |
| 2 | rs147749419 | CXCR1 | Regulatory | 0.017 | 0.154 | -0.043 | 2.94E-08 | Cognition decline rate |
| 8 | rs11787066 | LOC 107986930 | Intron | 0.148 | 0.276 | 0.015 | 6.93E-08 | β-Amyloid |
| 19 | rs34134669 | ADAMTS10 | Regulatory | 0.234 | 0.119 | -0.005 | 8.57E-07 | Cognition decline rate |
| 19 | rs769449 | APOE TOMM40 | Regulatory | 0.111 | 0.121 | 0.076 | 3.45E-11 | Alzheimer's Dementia |
|  |  |  |  | 0.112 | 0.116 | 0.022 | 1.51E-16 | Tangle density |
|  |  |  |  | 0.109 | 0.475 | -0.025 | 2.09E-15 | Cognition decline rate |
| 19 | rs429358 | APOE | Missense | 0.138 | 0.144 | 0.037 | 7.72E-13 | Alzheimer's Dementia |
|  |  |  |  | 0.138 | 0.631 | 0.037 | 1.17E-20 | Tangle density |
| | | | | 0.138 | 0.999 | 0.083 | 6.60E-27 | β-Amyloid |
|  |  |  |  | 0.139 | 0.999 | 0.089 | 1.19E-33 | Global AD pathology |
|  |  |  |  | 0.136 | 0.17 | -0.036 | 1.29E-17 | Cognition decline rate |
| 19 | rs7412 | APOE | Missense | 0.077 | 0.108 | -0.027 | 6.67E-13 | Global AD pathology |
| 19 | rs1065853 | APOC1 | Intergenic | 0.076 | 0.381 | -0.026 | 8.31E-13 | Global AD pathology |
| 19 | rs10414043 | APOC1 | Intergenic | 0.113 | 0.111 | 0.028 | 2.71E-12 | Alzheimer's Dementia |
| 19 | rs7256200 | APOC1 | Regulatory | 0.113 | 0.315 | 0.028 | 2.71E-12 | Alzheimer's Dementia |
|  |  |  |  | 0.113 | 0.228 | 0.03 | 3.86E-17 | Tangle density |
|  |  |  |  | 0.111 | 0.270 | -0.024 | 3.66E-15 | Cognition decline rate |
| 20 | rs1131695 | APOC1 | Stop gained | 0.435 | 0.119 | 0.039 | 1.06E-06 | Tangle density |

**Figure 3.**
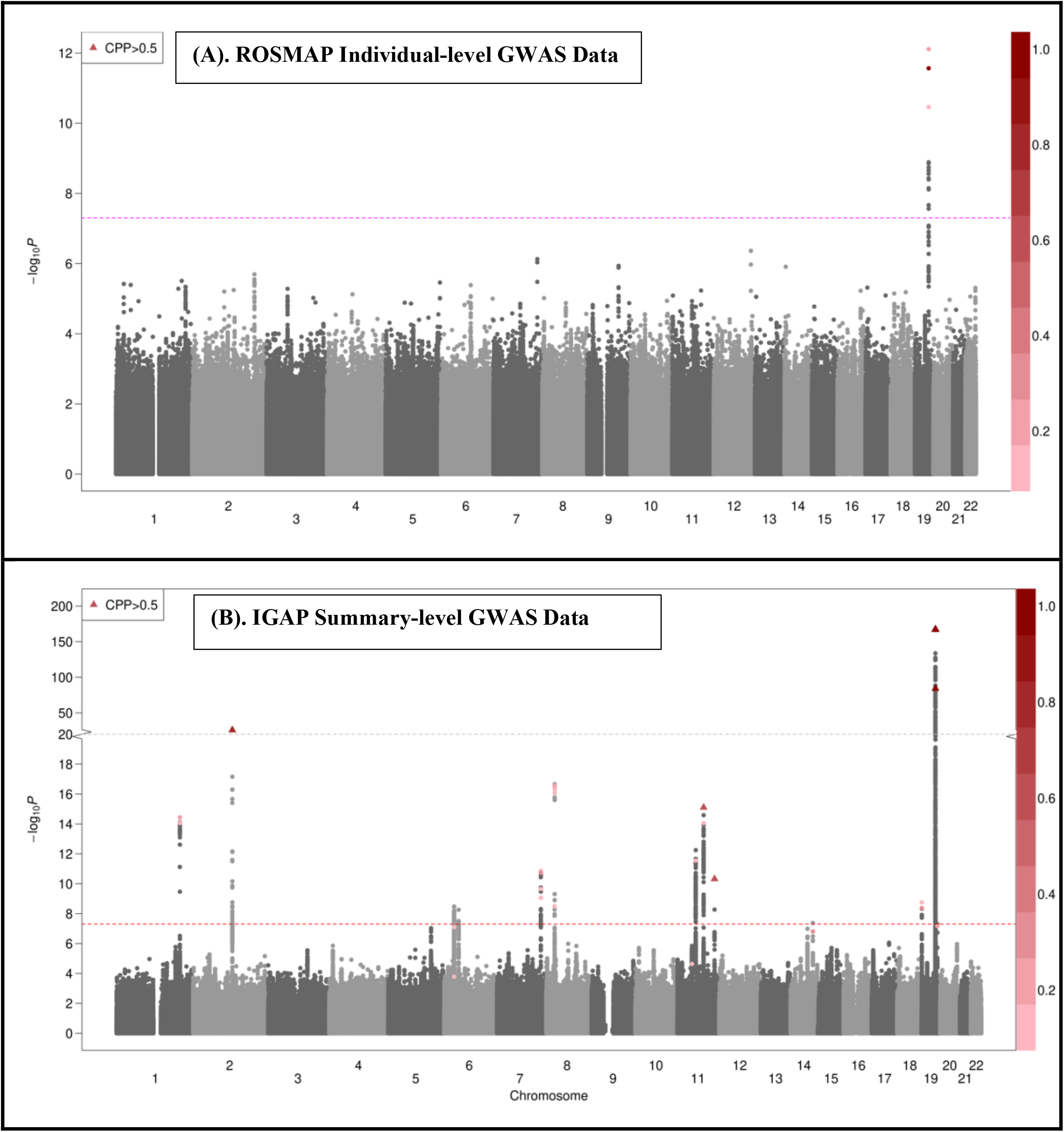
Manhattan plots of BFGWAS_QUANT results for studying Alzheimer’s dementia using ROS/MAP individual-level (A) and IGAP summary-level (B) GWAS data. Single variant test p-values were plotted in -log10 scale in the y-axis. The dashed horizontal line denotes the genome-wide significant threshold 5 **×** 10^−8^. SNPs with Bayesian CPP > 0.1068 were colored according to the color scale of their Bayesian CPP values by BFGWAS_QUANT. SNPs with Bayesian CPP >0.5 were plotted in solid triangles.

Using the IGAP summary-level GWAS data, BFGWAS_QUANT fine-mapped 32 significant SNPs with Bayesian CPP > 0.1068 associated with AD (Table 2; Figure 3B). Multiple SNPs located in genes *PVRL2, APOE/TOMM40, APOC1* in chromosome 19 were found to be associated with AD with CPP = 1. Interestingly, 10 out of 32 significant SNPs are cis-eQTL of Microglia, and all significant SNPs except one intergenic SNP (*rs7110631*) are intron, regulatory, downstream, upstream, 3’ UTR, and missense variants (Table 2). All significant SNPs except *rs78959900* are located in the peak regions of histone modification H3K27me3 (polycomb regression) that has the highest enrichment. Further, several SNPs that did not pass the genome-wide significance threshold (P-value < 5 × 10^−8^) were identified by integrating 10 functional annotations in BFGWAS_QUANT. These SNPs are located in genes that were previously found to be genetically linked to AD, such as *HLA-DRB1, NUP160, SLC24A4* and *CD33*^47; 58; 62-65^.

**Table 2.** Significant SNPs with Bayesian CPP > 0. 1068 by BFGWAS_QUANT for studying AD using the IGAP summary-level GWAS data. SNPs with single variant test P-value <5 × 10^−8^ were shaded in gray.

| CHR | rsID | Gene | Function | CPP | Beta | P-value |
| --- | --- | --- | --- | --- | --- | --- |
| 1 | rs6656401 | CR1 | Intron | 0.119 | -0.017 | 8.67E-15 |
| 1 | rs7515905 | CR1 | Intron | 0.206 | -0.019 | 3.75E-15 |
| 1 | rs1752684 | CR1 | Regulatory | 0.125 | -0.017 | 3.77E-15 |
| 1 | rs679515 | CR1 | Intron | 0.220 | -0.018 | 3.60E-15 |
| 2 | rs4663105 | BIN1 | Regulatory | 0.631 | 0.050 | 1.26E-26 |
| 2 | rs6733839 | BIN1 | Regulatory | 0.796 | 0.053 | 1.24E-26 |
| 6 | rs9270999 | HLA-DRB1 | Intron | 0.181 | 0.001 | 8.04E-08 |
| 6 | rs9273472 | HLA-DRB1 | Intron | 0.110 | 0.074 | 1.63E-04 |
| 7 | rs10808026 | EPHA1 | Intron | 0.123 | -0.020 | 1.36E-11 |
| 7 | rs11762262 | EPHA1 | Intron | 0.117 | -0.011 | 2.21E-10 |
| 7 | rs11763230 | EPHA1 | Intron | 0.325 | -0.020 | 1.86E-11 |
| 7 | rs11771145 | EPHA1 | Intron | 0.173 | -0.021 | 8.69E-10 |
| 8 | rs28834970 | PTK2B | Intron | 0.137 | 0.066 | 3.22E-09 |
| 8 | rs2279590 | CLU | Intron | 0.166 | 0.021 | 4.47E-17 |
| 8 | rs4236673 | CLU | Intron | 0.123 | 0.020 | 3.25E-17 |
| 8 | rs11787077 | CLU | Intron | 0.247 | 0.022 | 2.94E-17 |
| 8 | rs9331896 | CLU | Intron | 0.154 | 0.022 | 8.38E-17 |
| 8 | rs2070926 | CLU | Intron | 0.278 | 0.023 | 2.69E-17 |
| 11 | rs11039390 | NUP160 | Downstream | 0.145 | -0.004 | 2.31E-05 |
| 11 | rs4939338 | MS4A6E | Upstream | 0.139 | 0.011 | 2.79E-12 |
| 11 | rs7110631 | PICALM | Intergenic | 0.134 | 0.014 | 8.77E-15 |
| 11 | rs10792832 | RNU6-560P | Regulatory | 0.633 | 0.027 | 7.89E-16 |
| 11 | rs11218343 | SORL1 | Regulatory | 0.643 | -0.046 | 4.77E-11 |
| 14 | rs10498633 | SLC24A4 | Intron | 0.371 | -0.059 | 1.55E-07 |
| 19 | rs3752246 | ABCA7 | Missense | 0.361 | -0.027 | 4.27E-09 |
| 19 | rs4147929 | ABCA7 | Regulatory | 0.111 | -0.030 | 1.77E-09 |
| 19 | rs41289512 | PVRL2 | Regulatory | 1.000 | 0.132 | 1.81E-167 |
| 19 | rs6857 | PVRL2 | 3' UTR | 1.000 | 0.359 | 0 |
| 19 | rs769449 | APOE/TOMM40 | Regulatory | 1.000 | 0.292 | 0 |
| 19 | rs56131196 | APOC1 | Regulatory | 1.000 | 0.251 | 0 |
| 19 | rs78959900 | APOC1 | Downstream | 1.000 | -0.096 | 8.22E-85 |
| 19 | rs12459419 | CD33 | Missense | 0.245 | -0.027 | 6.66E-08 |

Additionally, the summation of genome-wide Bayesian CPP of SNPs with CPP >0.01 can be used to estimate the number of total causal SNPs for the phenotype of interest. The threshold of CPP>0.01 is used to exclude adding CPP from random MCMC selections. Although the power is limited for analyzing the ROS/MAP individual-level GWAS data with a small sample size, BFGWAS_QUANT estimated a total of 54 potential causal SNPs for AD using the IGAP summary-level GWAS data (Table 3).

**Table 3.** Estimates of total causal SNPs. The summations of the Bayesian CPP estimates of SNPs with CPP>0.01 estimate the total number of causal SNPs.

| GWAS Data | Phenotype | BFGWAS_QUANT | BVSR<sup>a</sup> |
| --- | --- | --- | --- |
| ROS/MAP | Alzheimer's dementia | 0.718 | 6.472 |
|  | Tangle density | 3.179 | 6.127 |
| | β-Amyloid | 5.375 | 7.316 |
|  | Global AD pathology | 5.375 | 6.174 |
|  | Cognition decline rate | 6.219 | 7.136 |
| IGAP | Alzheimer's dementia | 54.282 | - |

<sup>a</sup>. BVSR was not developed for using summary-level GWAS data.

### AD risk prediction by PRS in MCADGS

To show the usefulness of BFGWAS_QUANT for studying complex traits and diseases, we derived PRS using the Bayesian effect size estimates by BFGWAS_QUANT for an independent GWAS cohort MCADGS (n = 2099), and compared the risk prediction accuracy with the PRS using effect size estimates by BVSR, P+T, LDpred2, and PRS-CS. When the ROS/MAP individual-level GWAS data were used as training data, comparable AUC was obtained by using Bayesian effect size estimates byBFGWAS_QUANT (0.69) and BVSR (0.68), which was similar to the one obtained by LDpred2-auto (0.68) but significantly higher than the ones obtained by P+T (0.55), LDpred2-inf (0.53), and PRS-CS (0.54) (Figure 4A). This showed the advantage of deriving PRS using Bayesian effect size estimates by BFGWAS_QUANT when individual-level GWAS data are available, especially when the sample size is small. When the IGAP summary-level GWAS data were used as training data, the AUC by BFGWAS_QUANT (0.75) was the same as by LDpred2-auto (0.75), but much lower than P + T (0.88 with p-value threshold 10^−3^), LDpred2-inf (0.94), and PRS-CS (0.93) (Figure 4B). These results show that an infinitesimal model^66^ as assumed by PRS-CS and LDpred2-inf is more suitable for PRS development than the sparse model assumed by BFGWAS_QUANT and LDpred2-auto.

**Figure 4.**
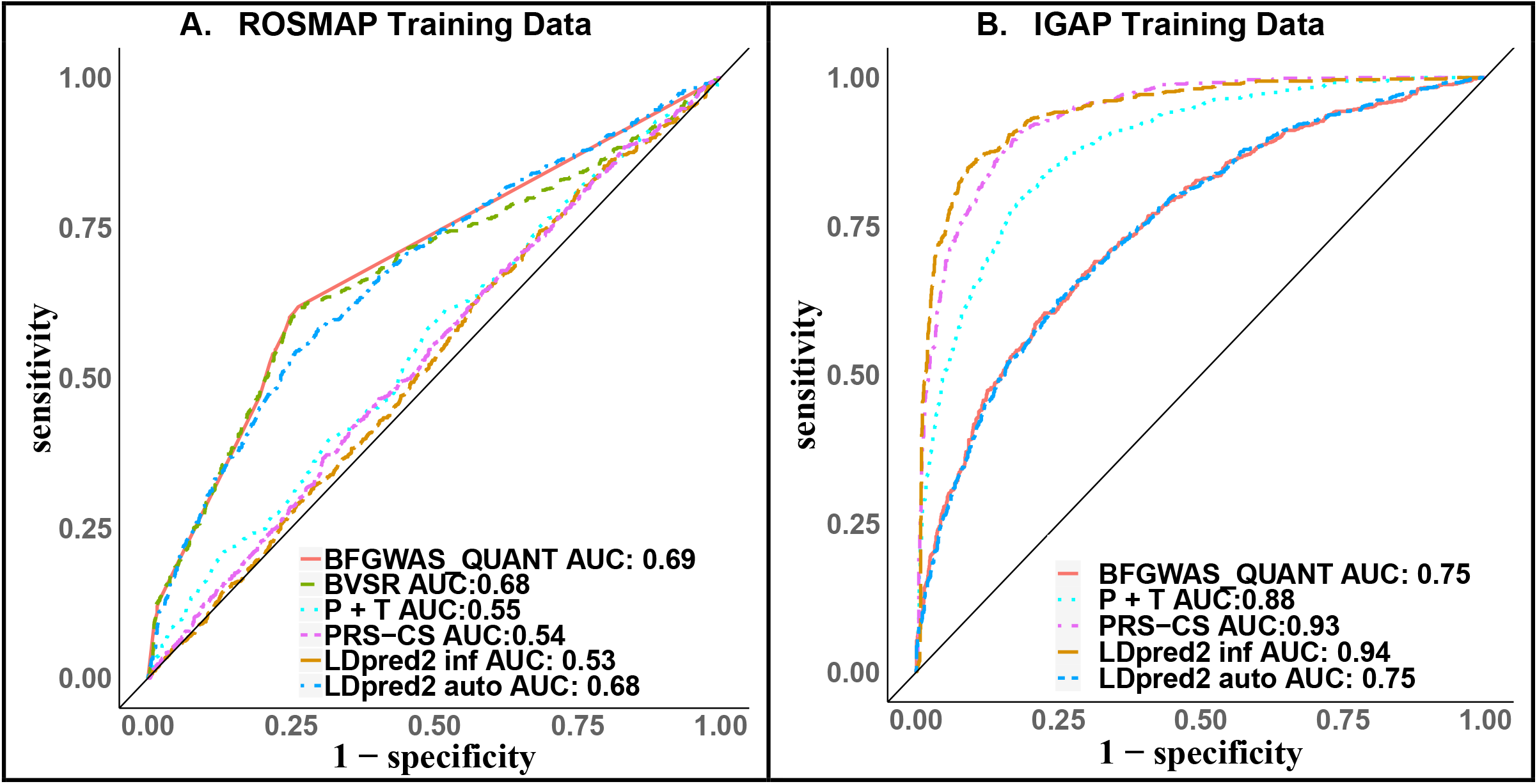
ROC plots comparing prediction accuracy of Alzheimer’s dementia in the test data of MCDGC by PRSs derived using the ROS/MAP individual-level (A) and IGAP summary-level (B) GWAS data as training data. PRS derived using Bayesian effect size estimates by BFGWAS_QUANT has comparable prediction accuracy as the PRSs derived by BVSR and LDpred2 auto, for all assuming a sparse causal model. PRSs derived by PRS-CS and LDpred2-inf using IGAP summary-level GWAS data as training data have the highest prediction accuracy for assuming an infinitesimal causal model.

## Discussion

We developed a scalable Bayesian functional GWAS method to account for multivariate quantitative functional annotations (BFGWAS_QUANT) for studying complex traits and diseases, which is based on a Bayesian hierarchical variable selection regression model and accompanied with a scalable EM-MCMC computation algorithm. BFGWAS_QUANT has the advantages of quantifying enrichment of functional annotations, as well as modeling LD to generate fine-mapped GWAS results that are also prioritized based on their functional annotations. BFGWAS_QUANT can be applied to both individual-level and summary-level GWAS data. In particular, the Bayesian effect size estimates can be used to derive PRS that accounts for functional annotations.

Our simulation studies validated the performance of BFGWAS_QUANT with respect to annotation enrichment quantification, GWAS association identification, and heritability estimation. Compared to BVSR, BFGWAS_QUANT method had higher sensitivities (i.e., power), while comparable PPV and accuracy of phenotype heritability estimation.

In the real studies of AD related phenotypes using both ROS/MAP individual-level and IGAP summary-level GWAS data, we showed that interesting enrichment patterns were identified, fine-mapped GWAS signals were identified, and predictive PRSs were derived. In particular, we found the histone modification H3K27me3 (polycomb regression) and Microglia cis-eQTL annotations were most enriched for association signals of AD. We also showed that SNPs with single variant test p-values < 5 **×** 10^−8^ could be identified for being prioritized due to their functional annotations.

Despite the above advantages, BFGWAS_QUANT does have its limitations. First, BFGWAS_QUANT model is developed for quantitative traits. However, following previous studies^22; 67^, GWAS analysis can still be done for dichotomous traits by quantifying cases as 1’s and controls as 0’s, which will have similar performance as a probit model if samples are independent and population structure can be addressed by top genotype principal components. Extending the BFGWAS_QUANT method for studying dichotomous traits is also part of our ongoing research. Second, BFGWAS_QUANT assumes the summary-level GWAS data and reference LD were derived from populations of the same ancestry. One need to apply BFGWAS_QUANT per ancestry first and then meta-analyze the results for studying GWAS cohorts with multiple ancestries. Third, BFGWAS_QUANT assumes a sparse causal genetic architecture which is suitable for generating fine-mapped GWAS results but might lack of power for deriving PRS.

In conclusion, our work demonstrated the usefulness of integrating multivariate quantitative functional annotations in GWAS for quantifying the enrichment of multiple functional annotations and generating fine-mapped GWAS results with higher power. Specifically, accurate quantification of annotation enrichment would help prioritize GWAS signals (fine-mapping) and then help illustrate the underlying genomic etiology of complex traits and diseases. Especially, as publicly available molecular QTL based and epigenomic features functional annotations continue to grow, BFGWAS_QUANT provides a convenient tool for integrative multi-omics studies.

## Data Availability

All data produced in the present study are available upon reasonable request to the authors

https://www.radc.rush.edu/

## Supplemental Data

Supplemental Note includes 9 Supplemental Figures and Supplemental Methods with details of the model derivations.

## Declaration of Interests

The authors declare no competing interests. David A. Bennett receives consulting fee from B4X inc for advising on neurodegenerative disease.

## Acknowledgements

J.Y. is supported by NIH/NIGMS grant R35GM138313 and NIH/NIA grant R21AG070659. ROS/MAP study data were provided by the Rush Alzheimer’s Disease Center, Rush University Medical Center, Chicago, IL. Data collection was supported through funding by NIA grants P30AG10161, R01AG15819, R01AG17917, R01AG30146, R01AG36836, U01AG32984, U01AG46152, U01AG61356, the Illinois Department of Public Health, and the Translational Genomics Research Institute. The MCADGC led by Dr. Nilüfer Ertekin-Taner and Dr. Steven G. Younkin, Mayo Clinic, Jacksonville, FL uses samples from the Mayo Clinic Study of Aging, the Mayo Clinic Alzheimer’s Disease Research Center, and the Mayo Clinic Brain Bank. MCADGC data collection was supported through funding by NIA grants P50 AG016574, R01 AG032990, U01 AG046139, R01 AG018023, U01 AG006576, U01 AG006786, R01 AG025711, R01 AG017216, R01 AG003949, NINDS grant R01 NS080820, CurePSP Foundation, and support from Mayo Foundation.

## Web Resources

BFGWAS_QUANT Github directory, https://github.com/yanglab-emory/BFGWAS_QUANT

RADC Research Resource Sharing Hub, http://www.radc.rush.edu/

ROS/MAP data, https://www.synapse.org/#!Synapse:syn3219045

MCADGC data, https://www.synapse.org/#!Synapse:syn2910256

LDpred, https://github.com/bvilhjal/ldpred

PRS-CS, https://github.com/getian107/PRScs

## Data and Code Availability

ROS/MAP data can be requested through Rush Alzheimer’s Disease Center (http://www.radc.rush.edu/) and synapse (https://www.synapse.org/#!Synapse:syn3219045). MCADGS data can be requested through synapse (https://www.synapse.org/#!Synapse:syn2910256). IGAP summary statistics are available from http://web.pasteur-lille.fr/en/recherche/u744/igap/igap_download.php. Source code of BFGWAS_QUANT is available through Github, https://github.com/yanglab-emory/BFGWAS_QUANT.

## Notes

### Competing Interest Statement

The authors have declared no competing interest.

### Funding Statement

NIGMS/NIA

