## Supplemental Data for "Scalable Bayesian functional GWAS method accounting for multivariate quantitative functional annotations with applications to studying Alzheimer’s disease"

#### Supplemental Figures

**Supplemental Figure 1. Bayesian enrichment estimates by BFGWAS_QUANT in simulation studies with phenotype heritability $h^2 = 0.5$, $\alpha_0 = (-10.5, -9.5)$.** Boxplots were made from Bayesian enrichment estimates from 100 replicated simulations with red dots denoting true enrichment values.

**A.** $\alpha_0 = -10.5$ | **B.** $\alpha_0 = -9.5$

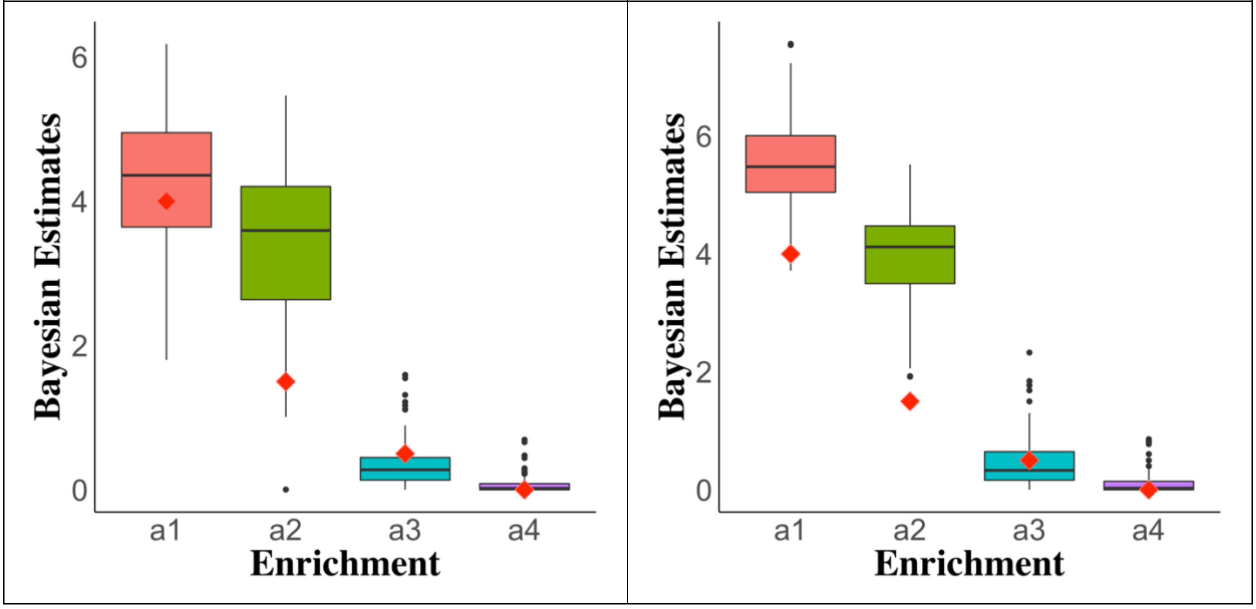

**Supplemental Figure 2. Positive predictive values (PPV) in 100 replicated simulations with $\alpha_0 = (-10.5, -9.5)$ by BFGWAS_QUAN (red) and BVSR (blue).**

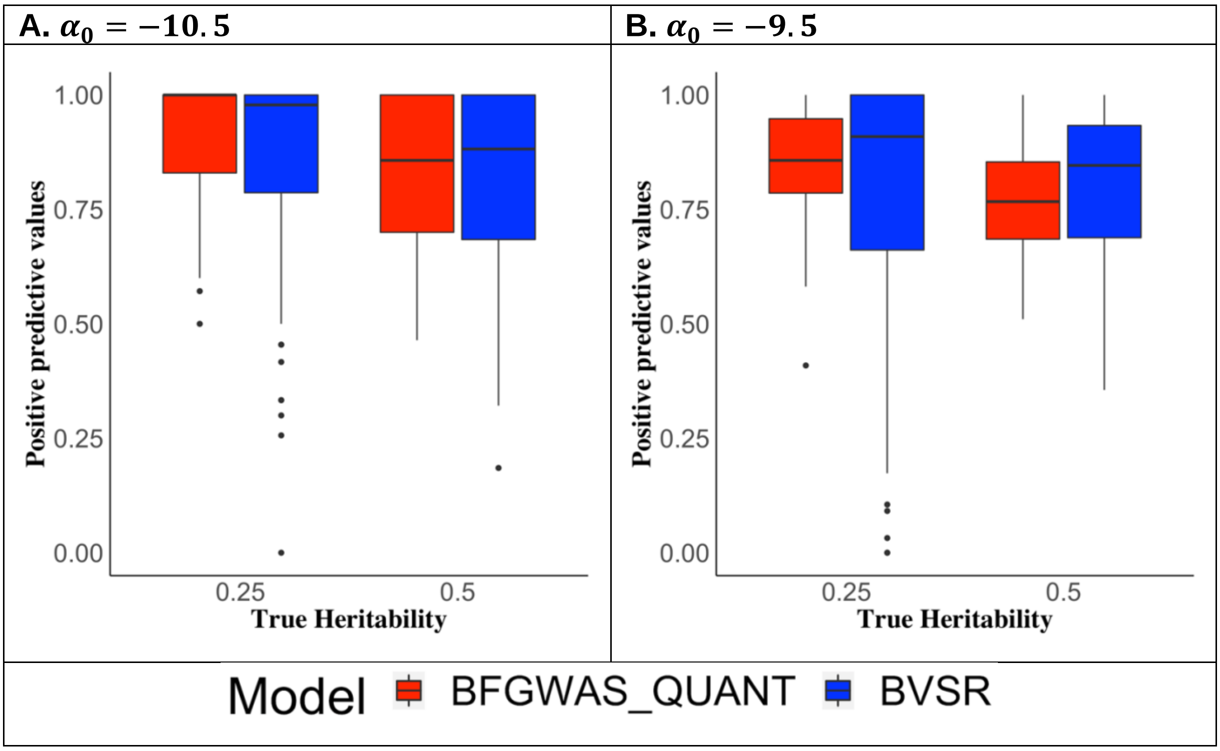

**Supplement Figure 3. Simulation results in the scenario where 10 true causal SNPs were randomly selected.**

**A. Bayesian enrichment estimates**

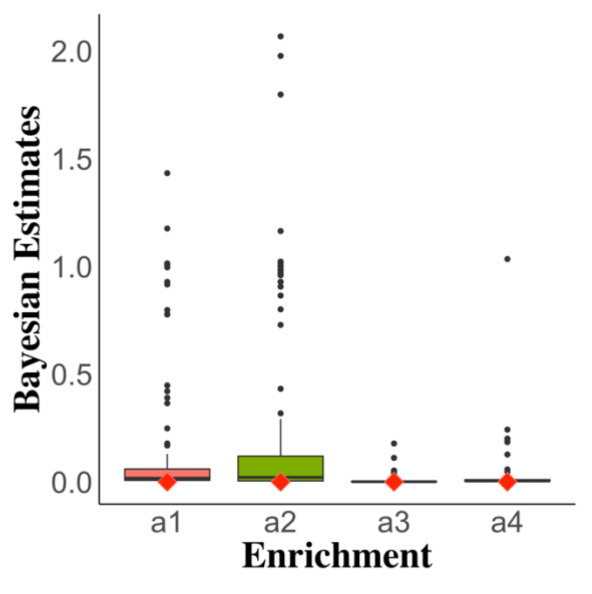


**B. Heritability**

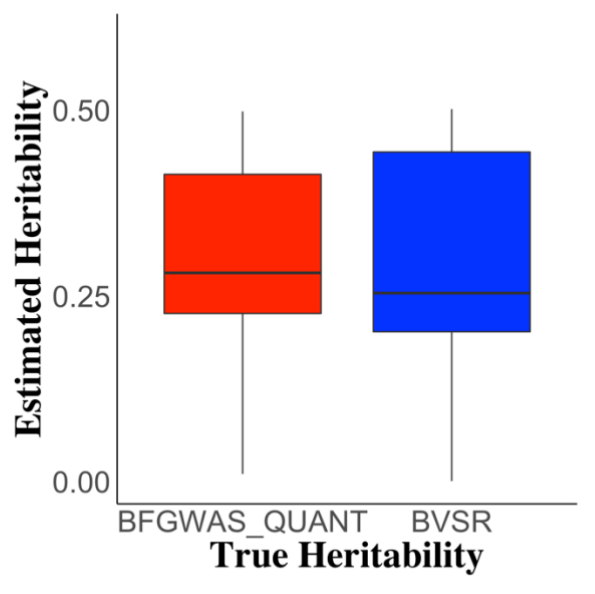


**C. Sensitivity**

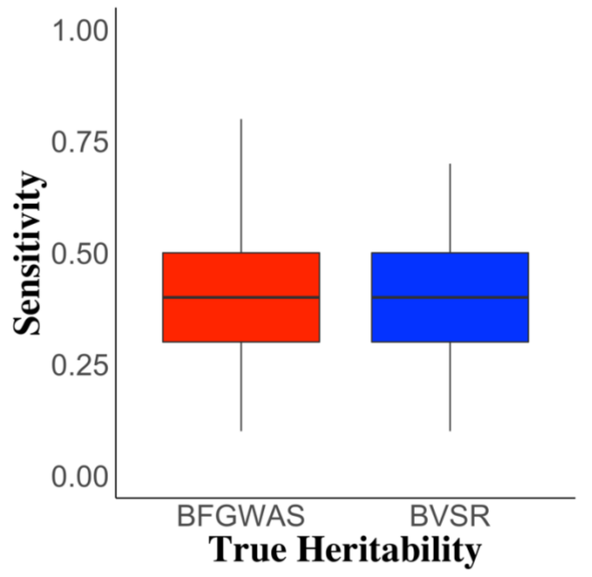


**D. Positive predictive value**

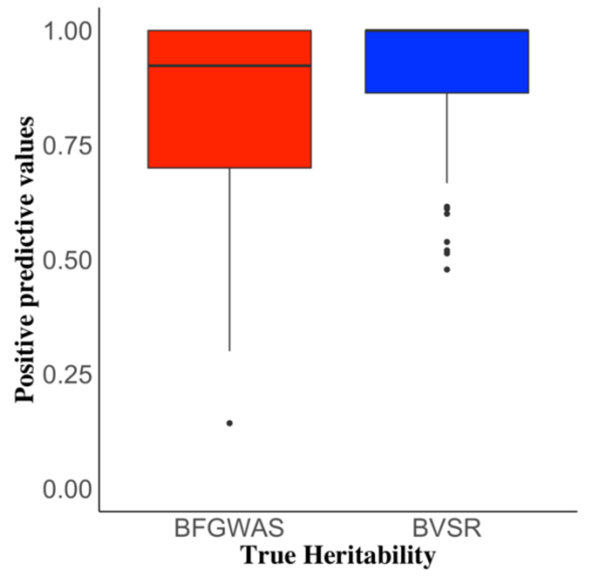

**Supplemental Figure 4. Manhattan plots for example simulations with $\alpha_0 = -10.5$ and heritability $h^2 = (0.25, 0.5)$.** SNPs with Bayesian CPP > 0.1068 were colored according to the color scale of CPP values. Red triangles denote true causal SNPs identified by BFGWAS_QUANT with CPP > 0.1068, and blue triangles denote true causal SNPs missed by BFGWAS_QUANT with CPP < 0.1068.

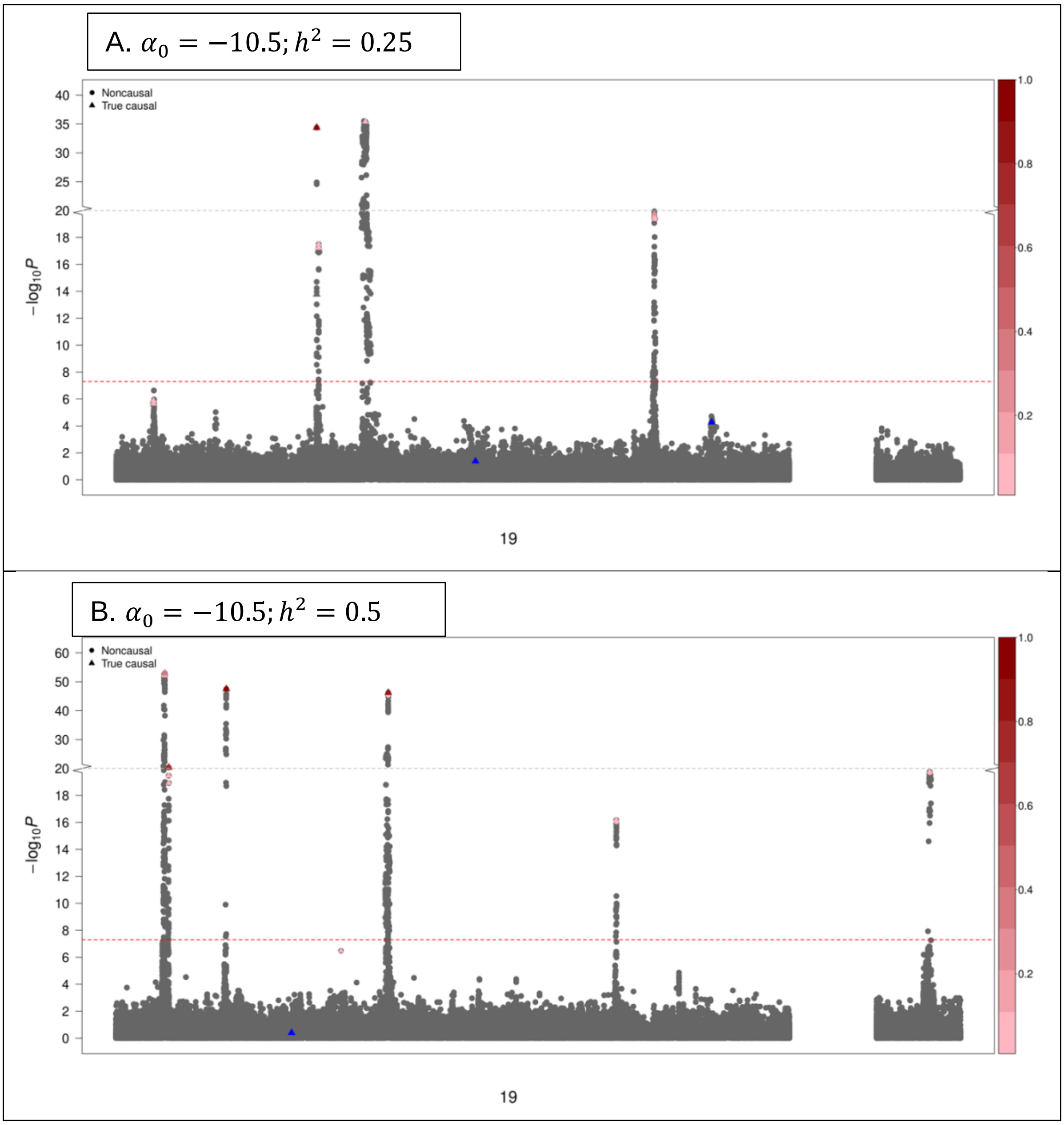

**Supplemental Figure 5. Manhattan plots for example simulations with $\alpha_0 = -9.5$ and heritability $h^2 = (0.25, 0.5)$.** SNPs with Bayesian CPP > 0.1068 were colored according to the color scale of CPP values. Red triangles denote true causal SNPs identified by BFGWAS_QUANT with CPP > 0.1068, and blue triangles denote true causal SNPs missed by BFGWAS_QUANT with CPP < 0.1068.

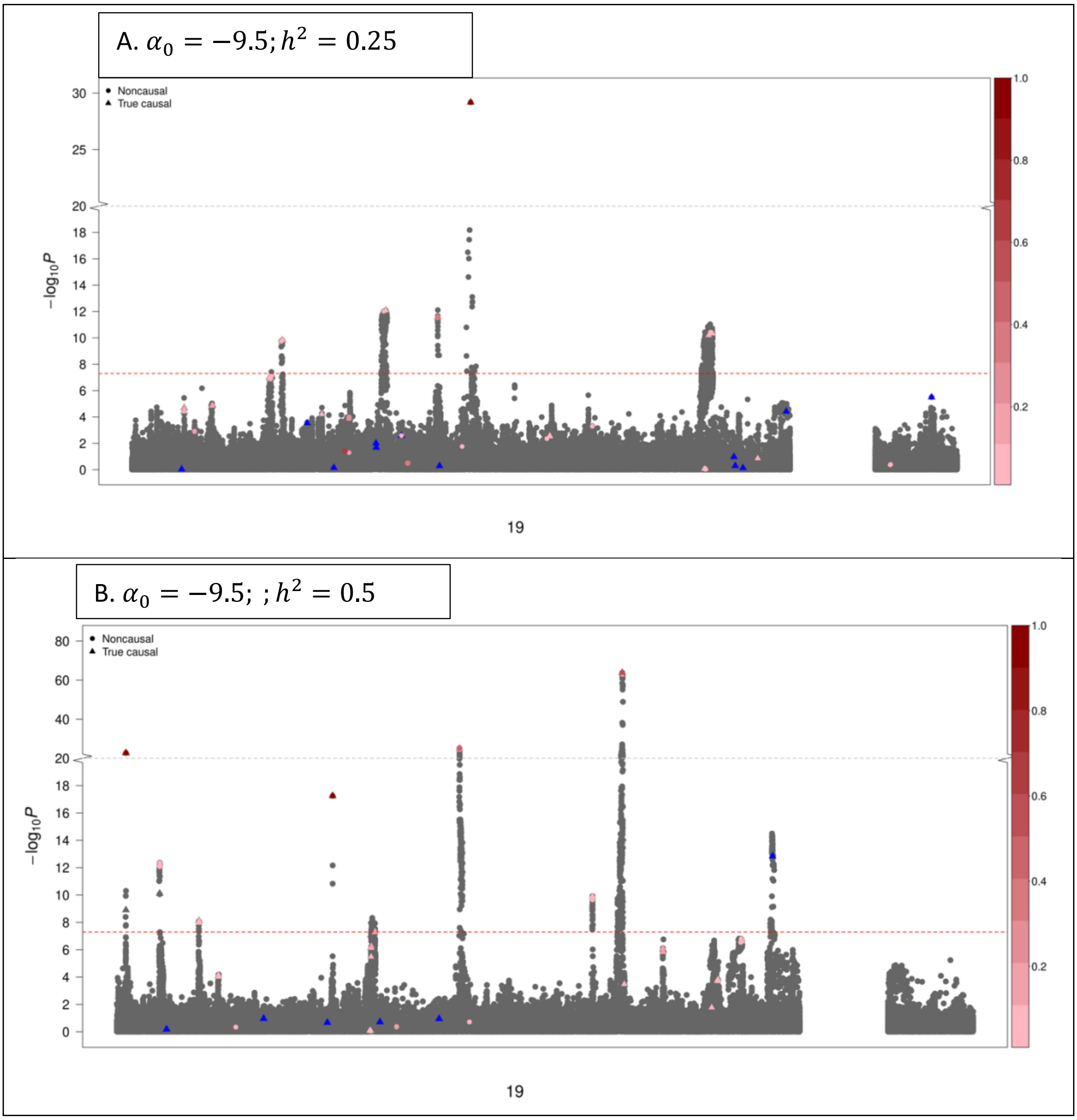

**Supplement Figure 6. Bayesian estimates of functional annotation enrichment for studying Tangle density (A) and $\beta$-Amyloid (B), using ROS/MAP individual-level GWAS data**.

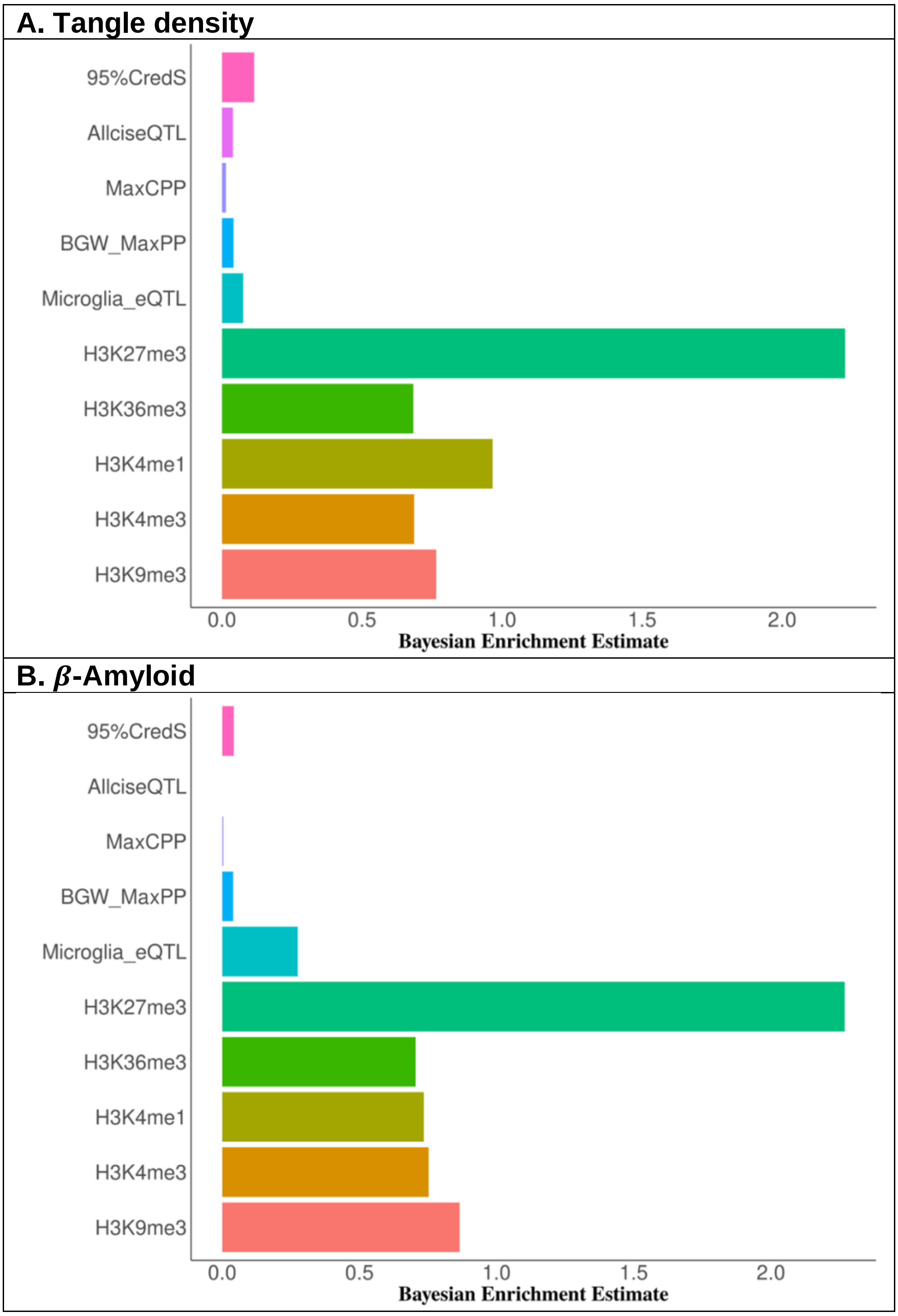

**Supplement Figure 7. Bayesian estimates of functional annotation enrichment for studying Global AD pathology (A) and Cognition decline rate (B), using ROS/MAP individual-level GWAS data.**

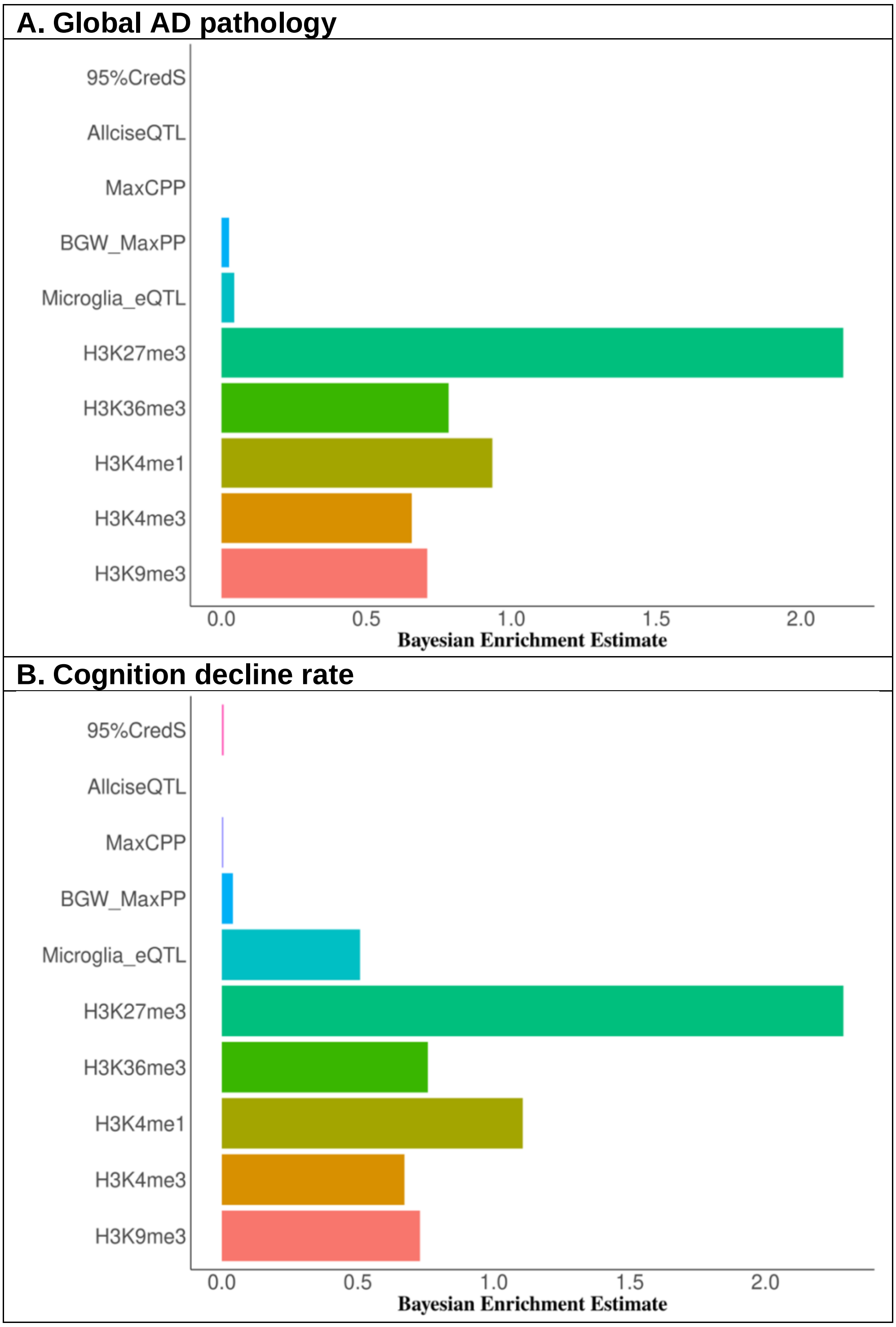

**Supplemental Figure 8. Manhattan plots of BFGWAS_QUANT results for Tangle density (A) and $\beta$-Amyloid (B), using ROS/MAP individual-level GWAS data.** Single variant test p-values were plotted in -log10 scale in the y-axis. The dashed horizontal line denotes the genome-wide significant threshold 5E-8. SNPs with Bayesian CPP > 0.1068 were colored according to the color scale of their Bayesian CPP values by BFGWAS_QUANT. SNPs with Bayesian CPP >0.5 were plotted in solid triangles.

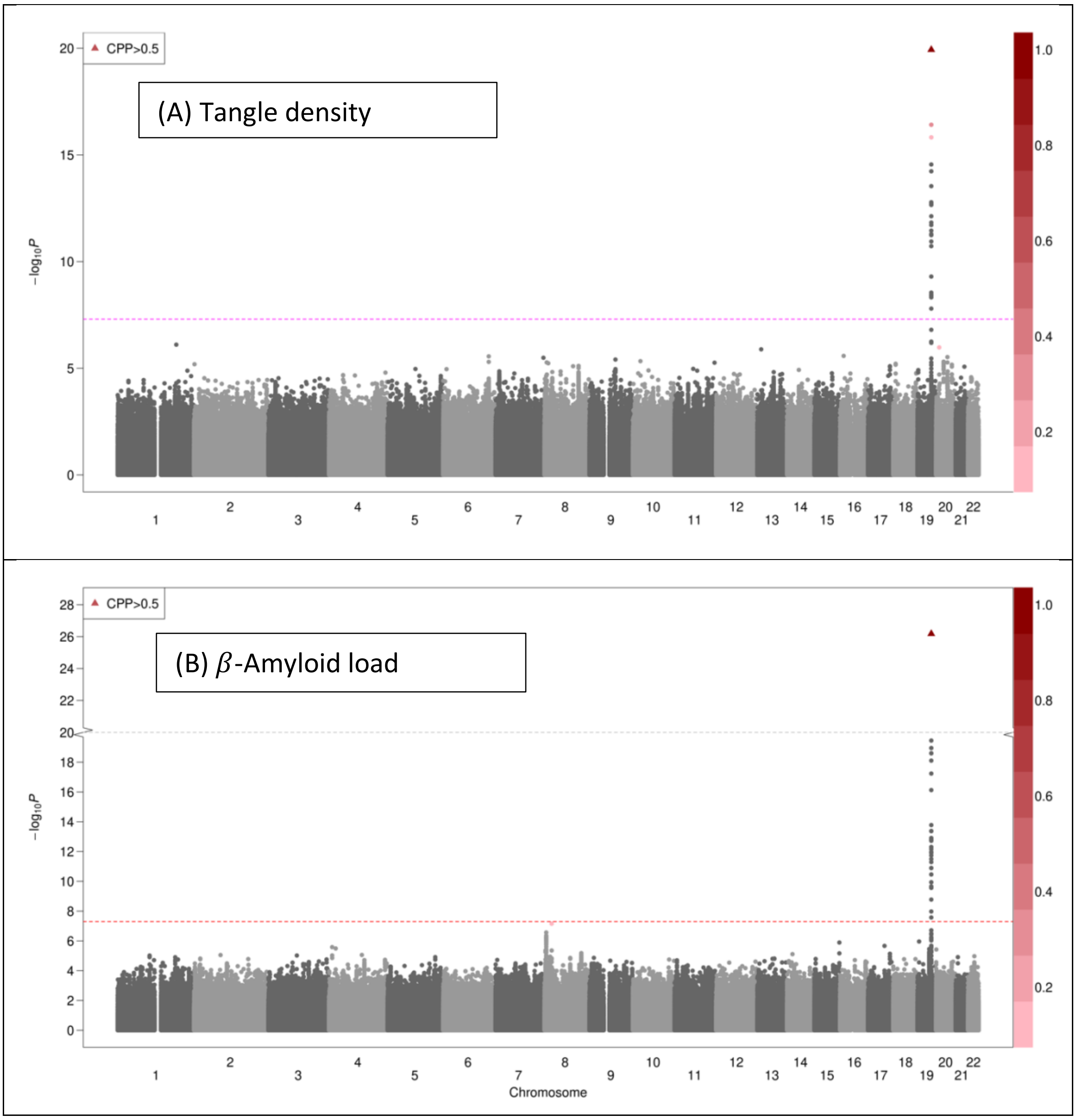

**Supplemental Figure 9. Manhattan plots of BFGWAS_QUANT results for Global AD pathology (A) and Cognition decline rate (B), using ROS/MAP individual-level GWAS data.** Single variant test p-values were plotted in -log10 scale in the y-axis. The dashed horizontal line denotes the genome-wide significant threshold $5 \times 10^{-8}$. SNPs with Bayesian CPP > 0.1068 were colored according to the color scale of their Bayesian CPP values by BFGWAS_QUANT. SNPs with Bayesian CPP >0.5 were plotted in solid triangles.

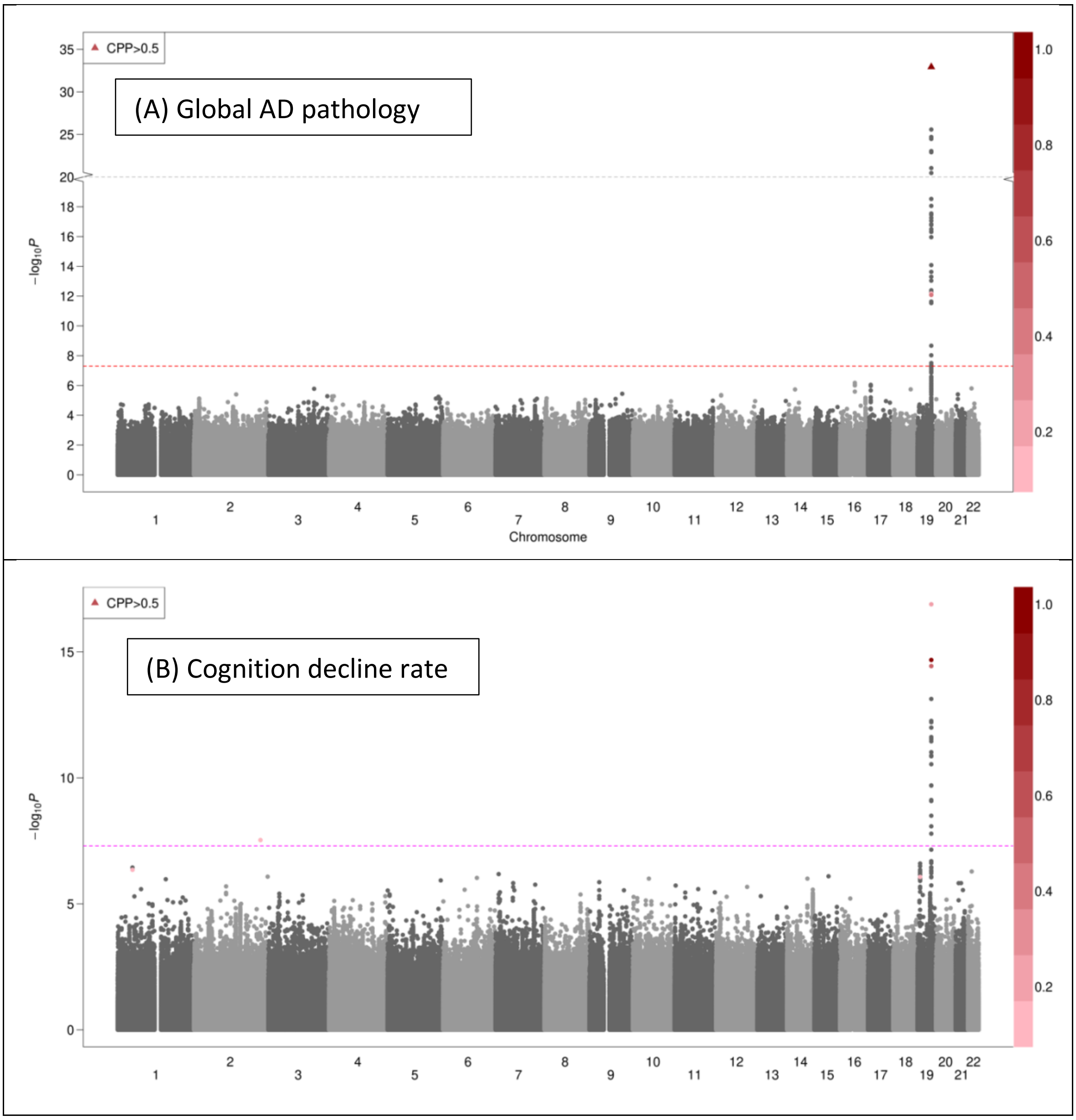

### Supplemental Methods

#### 1 Bayesian Hierarchical Variable Selection Regression Model

##### 1.1 Bayesian variable selection regression model

BFGWAS_QUANT assumes a multivariate Bayesian variable selection regression (BVSR) model for genome-wide variants [1]:

$$\boldsymbol{y}_{n\times 1} = \boldsymbol{X}_{n\times p}\boldsymbol{\beta}_{p\times 1} + \boldsymbol{\epsilon}_{n\times 1};\ \boldsymbol{\epsilon}_{n\times 1} \sim N(0, \boldsymbol{I}); \tag{1}$$

$$\beta_i \sim \pi_i N\left(0, \frac{1}{n}\tau_\beta^{-1}\right) + (1-\pi_i)\delta_0(\beta_i),\ i = 1, ..., p;$$

where $\boldsymbol{y}_{n\times 1}$ is a vector of standardized phenotype with $n$ samples; $\boldsymbol{X}_{n\times p}$ is a standardized genotype matrix with $p$ genetic variants; $\boldsymbol{\beta}_{p\times 1}$ is a vector of the genetic effect sizes; and $\boldsymbol{\epsilon}_{n\times 1}$ denotes the residuals which are independently distributed with a normal distribution, $N(0, \boldsymbol{I})$. We assume each $\beta_i$ follows a spike-and-slab prior distribution [2] – a mixture distribution of a normal distribution centered at zero and a point-mass density function at $0$, with $\delta_0(\beta_i) = 0$ if $\beta_i \neq 0$ and $\beta_i = 1$ otherwise. The assumed spike-and-slab prior for $\beta_i$ enforces variable selection in the regression model (1). Specifically, $\pi_i$ denotes the "casual" probability of variant $i$ for $\beta_i$ to be non-zero and following a normal distribution $N\left(0, \frac{1}{n}\tau_\beta^{-1}\right)$, and $(1-\pi_i)$ denotes the probability for $\beta_i = 0$.

We assume the genotype matrix $\boldsymbol{X}_{n\times p}$ is standardized from raw dosage data within range $[0, 2]$ or raw genotype data with values $\{0, 1, 2\}$, where each SNP column has mean $0$ and variance $1$. The intercept term is omitted from the regression model because both response and explanatory variables are standardized with mean $0$ and variance $1$. The residual variance is assume to be $1$ as equal to the standardized phenotype variance. The standardization of the phenotype vector and genotype matrix is important for using summary GWAS data.

We assume a scale of $\frac{1}{n}$ for the prior variance of $\beta_i$, for convenient derivation of the conditional posterior distributions. Hyper parameter $\tau_\beta$ will be given a prior fixed value in the domain of $(0, 1]$. With $\tau_\beta = 1$, $\beta_i$ is assumed to have the same prior variance as the marginal effect size estimate based on a single variant regression model. Larger values for $\tau_\beta$ will shrink posterior estimates for $\beta_i$ closer to $0$, while smaller values for $\tau_\beta$ might inflate the magnitude of posterior estimates for $\beta_i$. We recommend to select the prior $\tau_\beta$ value to ensure that the Bayesian posterior estimates are of similar magnitude as the marginal estimates.

Although BFGWAS_QUANT method is developed for quantitative traits, it can also be used for studying dichotomous trait by taking cases as $1$ and controls as $0$ when all samples are independent and the population structure can be addressed by top genotype principal components [3, 4].

##### 1.2 Account for multivariate quantitative functional annotations

We assume a hierarchical logistic model for causal probability $\pi_i$ in model 1 to account for multivariate quantitative functional annotations:

$$logit(\pi_i) = \boldsymbol{A_i'\alpha},\ i = 1, ..., p \tag{2}$$

where $\boldsymbol{A_i} = (1, A_{i1}, ..., A_{iJ})$ denotes the augmented annotation vector for the $i$th variant with an intercept term in first element, and $\boldsymbol{\alpha} = (\alpha_0, \alpha_1, ..., \alpha_J)'$ denotes the intercept coefficient $\alpha_0$ and corresponding enrichment coefficient $\{\alpha_j, j = (1, \cdots, J)\}$, with respect to the $j$th functional annotation.

We fix $\alpha_0$ as a value in the range of $[-13.8, -9]$ to assume a small prior "causal" probability for all genetic variants when $\{\alpha_j = 0,\ j = (1, \cdots, J)\}$. For example, $\alpha_0 = -13.8$ would assume prior "causal" probability $= 10^{-6}$ when $\{\alpha_j = 0,\ j = (1, \cdots, J)\}$. We assume a standard normal prior for enrichment coefficient $\{\alpha_j \sim N(0, 1);\ j = 1, ...J\}$.

##### 1.3 Latent Indicator Variable

To facilitate computation, a latent indicator vector $\boldsymbol{\gamma}_{p\times 1}$ is introduced for model 1, where each element $\gamma_i \in \{0, 1\}$ indicates whether the corresponding $i$th genetic effect $\beta_i$ equals to $0$ with $\gamma_i = 0$ or follows the $N(0, \sigma_\beta^2)$ distribution with $\gamma_i = 1$. Equivalently,

$$\gamma_i \sim Bernoulli(\pi_i),\ \boldsymbol{\beta_{-\gamma}} \sim \delta_0(\cdot),\ \boldsymbol{\beta_\gamma} \sim MVN_{|\boldsymbol{\gamma}|}\left(0, \frac{1}{n}\tau_\beta^{-1}\boldsymbol{I_\gamma}\right), \tag{3}$$

where $|\boldsymbol{\gamma}|$ denotes the number of non-zero entries in $\boldsymbol{\gamma}$; $\boldsymbol{\beta_{-\gamma}}$ denotes the sub-vector of $\boldsymbol{\beta}_{p\times 1}$ corresponding to variants with $\gamma_i = 0$; $\boldsymbol{\beta_\gamma}$ denotes the sub-vector of $\boldsymbol{\beta}_{p\times 1}$ corresponding to the variants with $\{\gamma_j = 1; j = 1, \cdots, |\boldsymbol{\gamma}|\}$ that follows a multivariate normal distribution (MVN) with mean $0$ and variance-covariance matrix $\frac{1}{n}\tau_\beta^{-1}\boldsymbol{I_\gamma}$; and $\boldsymbol{I}_\gamma$ is an $|\gamma| \times |\gamma|$ identity matrix. Then the expected value of this indicator, $E[\gamma_i]$ represents the causal posterior probability $(CPP_i)$ for each genetic variant to have a non-zero effect size (i.e., to be a causal genetic variant for the phenotype of interest).

##### 1.4 Bayesian inference

From the assumed BVSR model (1), the posterior joint distribution of parameters of interest, $(\boldsymbol{\beta}, \gamma, \boldsymbol{\pi}(\boldsymbol{\alpha}))$, is proportional to the product of likelihood and prior density functions,

$$P(\boldsymbol{\beta}, \boldsymbol{\gamma}, \boldsymbol{\pi}(\boldsymbol{\alpha})|\boldsymbol{y}, \boldsymbol{X}, \boldsymbol{A}) \propto P(\boldsymbol{y}|\boldsymbol{X}, \boldsymbol{\beta}, \gamma)P(\boldsymbol{\beta}|\gamma, \tau_\beta)P(\gamma|\boldsymbol{\pi}(\boldsymbol{\alpha}), \boldsymbol{A})P(\boldsymbol{\pi}(\boldsymbol{\alpha})). \tag{4}$$

Our main goal is to infer parameters of interest, including genetic effect sizes $\boldsymbol{\beta}$, expected causal probability $\boldsymbol{E}[\gamma]$, and annotation enrichment coefficients $\boldsymbol{\alpha}$, while conditioning on the data $(\boldsymbol{y}, \boldsymbol{X}, \boldsymbol{A})$. In order to make the Bayesian inference applicable in practice with millions of genetic variants, we employ the scalable Expectation-Maximization Markov chain Monte Carlo (EM-MCMC) algorithm as used by the original BFGWAS method, referred to as BFGWAS_CAT in this paper [1].

Specifically, we first segment considered genotype data into approximately independent genome-blocks based on the block-wise linkage disequilibrium (LD) structure of the human genome, i.e., $\boldsymbol{X} = \{\boldsymbol{X}_1, \boldsymbol{X}_2, \cdots, \boldsymbol{X}_K\}$. Then we can write the likelihood function (4) as a product of likelihood functions for $\boldsymbol{X}_k$,

$$P(\boldsymbol{y}|\boldsymbol{X}, \boldsymbol{\beta}, \gamma) = \prod_{k=1}^{K} P_k(\boldsymbol{y}|\boldsymbol{X}_k, \boldsymbol{\beta}_k, \gamma_k), \tag{5}$$

where $(\boldsymbol{y}|\boldsymbol{X}_k, \boldsymbol{\beta}_k, \gamma_k) \sim MVN_{|\gamma_{\boldsymbol{k}}|}(\boldsymbol{X}_k\boldsymbol{\beta}_k, \boldsymbol{I}_{|\gamma_{\boldsymbol{k}}|})$.

By EM-MCMC algorithm, given values of $(\boldsymbol{\alpha}, \tau_\beta)$, we estimate $(\boldsymbol{\beta_k}, \boldsymbol{E}[\gamma_{\boldsymbol{k}}])$ by implementing MCMC algorithm [5, 6] per genome-block (i.e., Expectation step (E-step)) where $\boldsymbol{E}[\gamma_{\boldsymbol{k}}]$ is the Bayesian CPP. Then we update the estimates of $\boldsymbol{\alpha}$ by maximizing the corresponding expected posterior likelihood function [7] (Maximization step (M-step)), given the genome-wide estimates of $(\boldsymbol{\beta}, \boldsymbol{E}[\gamma])$ from the previous E-step. A few such EM iterations will be run until the estimates of $\boldsymbol{\alpha}$ converge, which generally requires 3-5 EM iterations [1]. The derivation of the maximum a posterior probability (MAP) estimates for $\boldsymbol{\alpha}$ is described in Section 2.2.

###### 1.4.1 Conditional posterior distribution for genetic effect sizes

Conditioning on the values of $(\boldsymbol{\pi}(\boldsymbol{\alpha}), \tau_\beta)$, the posterior distribution for the variant-specific parameters $(\boldsymbol{\beta}, \gamma)$ is

$$P(\boldsymbol{\beta}, \boldsymbol{\gamma}|\boldsymbol{X}, \boldsymbol{y}, \boldsymbol{\pi}(\boldsymbol{\alpha}), \tau_\beta) \propto P(\boldsymbol{y}|\boldsymbol{X}, \boldsymbol{\beta}, \gamma)P(\boldsymbol{\beta}|\gamma, \tau_\beta)P(\boldsymbol{\gamma}|\boldsymbol{\pi}(\boldsymbol{\alpha})). \tag{6}$$

Conditioning on $\boldsymbol{\gamma}$, the effect-sizes associated with a $\gamma$ of zero equal 0, while the posterior distribution for m effect-sizes associated with a non-zero $\gamma$, denoted as $\boldsymbol{\beta}_{|\gamma|}$, is given by

$$P(\boldsymbol{\beta}_{|\gamma|}|\boldsymbol{y}, \boldsymbol{X}, \boldsymbol{\gamma}, \tau_\beta) \propto P(\boldsymbol{y}|\boldsymbol{X}, \boldsymbol{\beta}_{|\gamma|}, \boldsymbol{\gamma}) P(\boldsymbol{\beta}_{|\gamma|}|\boldsymbol{\gamma}, \tau_\beta)$$

$$\propto \exp\left[-\frac{1}{2}(\boldsymbol{y} - \boldsymbol{X}\boldsymbol{\beta}_{|\gamma|})^T(\boldsymbol{y} - \boldsymbol{X}\boldsymbol{\beta}_{|\gamma|})\right] \exp\left[-\frac{1}{2}\boldsymbol{\beta}_{|\gamma|}^T(n\tau_\beta \boldsymbol{I_{m\times m}})\boldsymbol{\beta}_{|\gamma|}\right]$$

$$\propto \exp\left\{-\frac{1}{2}\left[\boldsymbol{\beta}_{|\gamma|}^T(\boldsymbol{X}^T\boldsymbol{X} + n\tau_\beta \boldsymbol{I_{m\times m}})\boldsymbol{\beta}_{|\gamma|} - 2\boldsymbol{\beta}_{|\gamma|}^T\boldsymbol{X}^T\boldsymbol{y}\right]\right\}. \quad (7)$$

From (8), it is easy to see that

$$P(\boldsymbol{\beta}_{|\gamma|}|\boldsymbol{y}, \boldsymbol{X}, \boldsymbol{\gamma}, \tau_\beta) \sim MVN_{|\gamma|}(\boldsymbol{\mu}_{\boldsymbol{\beta}_{|\gamma|}}, \boldsymbol{\Sigma}_{\boldsymbol{\beta}_{|\gamma|}}), \quad (8)$$

where

$$\boldsymbol{\Sigma}_{\boldsymbol{\beta}_{|\gamma|}} = (\boldsymbol{X}^T\boldsymbol{X} + n\tau_\beta \boldsymbol{I_{m\times m}})^{-1} = \frac{1}{n}(\boldsymbol{R} + \tau_\beta \boldsymbol{I_{m\times m}})^{-1} = \frac{1}{n}\boldsymbol{\Omega^{-1}}, \quad (9)$$

$$\boldsymbol{R} = \frac{1}{n}\boldsymbol{X}^T\boldsymbol{X},\ \boldsymbol{\Omega} = \boldsymbol{R} + \tau_\beta \boldsymbol{I_{m\times m}};$$

and

$$\boldsymbol{\mu}_{\boldsymbol{\beta}_{|\gamma|}} = \boldsymbol{\Sigma}_{\boldsymbol{\beta}_{|\gamma|}}\boldsymbol{X}^T\boldsymbol{y}.$$

Since columns of $\boldsymbol{X}$ are standardized, $\boldsymbol{X}^T\boldsymbol{X} = n\boldsymbol{R}$ with $\boldsymbol{R}$ equal to the genotype correlation matrix (i.e., Reference LD).

Note that the genetic effect size based on a single variable regression model is given by, $\widehat{\beta}_j = \frac{\boldsymbol{X}_j^T\boldsymbol{y}}{n}$, with standardized genotype and phenotype vector. Then the $Z_{score}$ statistics are given by,

$$Z_{score_j} = \frac{\widehat{\beta}_j}{SE(\widehat{\beta}_j)} = \frac{\frac{1}{n}\boldsymbol{X_j^T y}}{\sqrt{Var(\boldsymbol{y})(\boldsymbol{X_j' X_j})^{-1}}} = \frac{\frac{1}{n}\boldsymbol{X_i^T y}}{\frac{1}{\sqrt{n}}} = \frac{\boldsymbol{X_j^T y}}{\sqrt{n}}.$$

Thus, given single variant test $\boldsymbol{Z_{score}}$ and sample size $n$ from GWAS summary data, the effect sizes can be derived by

$$\widehat{\beta}_j = \sqrt{n}Z_{score_j}.$$

That is,

$$\boldsymbol{\mu}_{\boldsymbol{\beta}_{|\gamma|}} = \boldsymbol{\Sigma}_{\boldsymbol{\beta}_{|\gamma|}}\boldsymbol{X^T y} = \boldsymbol{\Omega^{-1}}\widehat{\boldsymbol{\beta}}. \quad (10)$$

##### 1.4.2 Conditional posterior distribution for latent indicator variable

Because of the conditional conjugate prior for $\boldsymbol{\beta}$, we can easily integrate $\boldsymbol{\beta}$ out from the joint conditional posterior distribution (6) to obtain the marginal conditional posterior distribution for $\boldsymbol{\gamma}$,

$$P(\boldsymbol{\gamma}|\boldsymbol{X},\boldsymbol{y},\boldsymbol{\pi(\alpha)},\tau_\beta) \propto \int_{\boldsymbol{\beta}_{|\gamma|}} P(\boldsymbol{y}|\boldsymbol{X},\boldsymbol{\beta}_{|\gamma|},\tau_\beta)P(\boldsymbol{\beta}_{|\gamma|}|\boldsymbol{\gamma},\tau_\beta)d\boldsymbol{\beta}_{|\gamma|} \cdot P(\boldsymbol{\gamma}|\boldsymbol{\pi(\alpha)})$$

$$\propto P(\boldsymbol{\gamma}|\boldsymbol{\pi(\alpha)}) \cdot \int_{\boldsymbol{\beta}_{|\gamma|}} \frac{1}{\sqrt{(2\pi)^n}\sqrt{|\boldsymbol{I_n}|}} \exp\left\{-\frac{1}{2}(\boldsymbol{y}-\boldsymbol{X}\boldsymbol{\beta}_{|\gamma|})^T(\boldsymbol{y}-\boldsymbol{X}\boldsymbol{\beta}_{|\gamma|})\right\}$$
$$\cdot \frac{1}{\sqrt{(2\pi)^m}\sqrt{|\frac{1}{n\tau_\beta}\boldsymbol{I_m}|}} exp\left\{-\frac{1}{2}(\boldsymbol{\beta}_{|\gamma|}^T(n\tau_\beta\boldsymbol{I})\boldsymbol{\beta}_{|\gamma|})\right\} d\boldsymbol{\beta}_{|\gamma|}$$

$$\propto \frac{P(\boldsymbol{\gamma}|\boldsymbol{\pi(\alpha)})}{\sqrt{(2\pi)^n}\sqrt{(2\pi)^m}\sqrt{|\frac{1}{n\tau_\beta}\boldsymbol{I_m}|}} \cdot \int_{\boldsymbol{\beta}_{|\gamma|}} \exp\left\{-\frac{1}{2}\left[\boldsymbol{\beta}_{|\gamma|}^T(\boldsymbol{X}^T\boldsymbol{X}+n\tau_\beta\boldsymbol{I})\boldsymbol{\beta}_{|\gamma|} - 2\boldsymbol{\beta}_{|\gamma|}^T(\boldsymbol{X}^T\boldsymbol{y}) + \boldsymbol{y}^T\boldsymbol{y}\right]\right\}$$

$$\propto \frac{P(\boldsymbol{\gamma}|\boldsymbol{\pi(\alpha)})}{\sqrt{(2\pi)^m}\sqrt{|\frac{1}{n\tau_\beta}\boldsymbol{I_m}|}} \cdot \int_{\boldsymbol{\beta}_{|\gamma|}} \exp\{-\frac{1}{2}[\boldsymbol{\beta}_{|\gamma|}^T\boldsymbol{\Sigma}_{\boldsymbol{\beta}_{|\gamma|}}^{-1}\boldsymbol{\beta}_{|\gamma|} - 2\boldsymbol{\beta}_{|\gamma|}\boldsymbol{\Sigma}_{\boldsymbol{\beta}_{|\gamma|}}^{-1}\boldsymbol{\Sigma}_{\boldsymbol{\beta}_{|\gamma|}}(\boldsymbol{X}^T\boldsymbol{y})$$
$$+\boldsymbol{y}^T\boldsymbol{X}\boldsymbol{\Sigma}_{\boldsymbol{\beta}_{|\gamma|}}\boldsymbol{\Sigma}_{\boldsymbol{\beta}_{|\gamma|}}^{-1}\boldsymbol{\Sigma}_{\boldsymbol{\beta}_{|\gamma|}}\boldsymbol{X}^T\boldsymbol{y} + \boldsymbol{y}^T\boldsymbol{y} - \boldsymbol{y}^T\boldsymbol{X}\boldsymbol{\Sigma}_{\boldsymbol{\beta}_{|\gamma|}}\boldsymbol{\Sigma}_{\boldsymbol{\beta}_{|\gamma|}}^{-1}\boldsymbol{\Sigma}_{\boldsymbol{\beta}_{|\gamma|}}\boldsymbol{X}^T\boldsymbol{y}]\}d\boldsymbol{\beta}_{|\gamma|}$$

$$\propto \frac{P(\boldsymbol{\gamma}|\boldsymbol{\pi(\alpha)})\sqrt{|\boldsymbol{\Sigma}_{\boldsymbol{\beta}_{|\gamma|}}|}}{\sqrt{|\frac{1}{n\tau_\beta}\boldsymbol{I_m}|}} \cdot$$
$$\int_{\boldsymbol{\beta}_{|\gamma|}} \frac{1}{\sqrt{(2\pi)^m}\sqrt{|\boldsymbol{\Sigma}_{\boldsymbol{\beta}_{|\gamma|}}|}} \exp\left\{-\frac{1}{2}\left[(\boldsymbol{\beta}_{|\gamma|} - \boldsymbol{\Sigma}_{\boldsymbol{\beta}_{|\gamma|}}\boldsymbol{X}^T\boldsymbol{y})^T\boldsymbol{\Sigma}_{\boldsymbol{\beta}_{|\gamma|}}^{-1}(\boldsymbol{\beta}_{|\gamma|} - \boldsymbol{\Sigma}_{\boldsymbol{\beta}_{|\gamma|}}\boldsymbol{X}^T\boldsymbol{y})\right]\right\} d\boldsymbol{\beta}_{|\gamma|} \cdot$$
$$\exp\left\{-\frac{1}{2}\left[\boldsymbol{y}^T\boldsymbol{y} - \boldsymbol{y}^T\boldsymbol{X}\boldsymbol{\Sigma}_{\boldsymbol{\beta}_{|\gamma|}}\boldsymbol{\Sigma}_{\boldsymbol{\beta}_{|\gamma|}}^{-1}\boldsymbol{\Sigma}_{\boldsymbol{\beta}_{|\gamma|}}\boldsymbol{X}^T\boldsymbol{y}\right]\right\}$$

$$\propto \frac{\sqrt{|\boldsymbol{\Sigma}_{\boldsymbol{\beta}_{|\gamma|}}|}}{\sqrt{|\frac{1}{n\tau_\beta}\boldsymbol{I_m}|}} \cdot \exp\{-\frac{1}{2}\boldsymbol{y}^T\boldsymbol{y} + \frac{1}{2}\boldsymbol{y}^T\boldsymbol{X}\boldsymbol{\Sigma}_{\boldsymbol{\beta}_{|\gamma|}}\boldsymbol{X}^T\boldsymbol{y}\} \cdot P(\boldsymbol{\gamma}|\boldsymbol{\pi(\alpha)})$$

$$\propto \sqrt{|\boldsymbol{\Sigma}_{\boldsymbol{\beta}_{|\gamma|}}|} \cdot (n\tau_\beta)^{\frac{m}{2}} \cdot \exp\left\{-\frac{n}{2} + \frac{1}{2}(\boldsymbol{X}^T\boldsymbol{y})^T\boldsymbol{\Sigma}_{\boldsymbol{\beta}_{|\gamma|}}(\boldsymbol{X}^T\boldsymbol{y})\right\} \cdot \prod_{i=1}^{P} P(\gamma_i|\pi(\alpha_i)) \tag{11}$$

Log conditional posterior likelihood of $\gamma$ could be derived from (11),

$$
\begin{aligned}
& ln(P(\boldsymbol{\gamma}|\boldsymbol{X},\boldsymbol{y},\boldsymbol{\pi(\alpha)},\tau_\beta)) \\
&= C + \frac{1}{2}ln|\boldsymbol{\Sigma}_{\boldsymbol{\beta}_{|\gamma|}}| + \frac{m}{2}ln(n\tau_\beta) + \frac{1}{2}(\boldsymbol{X}^T\boldsymbol{y})^T\boldsymbol{\Sigma}_{\boldsymbol{\beta}_{|\gamma|}}(\boldsymbol{X}^T\boldsymbol{y}) + \sum_{i=1}^{P}[\gamma_i ln\pi(\alpha_i) + (1-\gamma_i)ln(1-\pi(\alpha_i)] \\
&= C + \frac{1}{2}ln|\frac{1}{n}\boldsymbol{\Omega^{-1}}| + \frac{m}{2}ln(n\tau_\beta) + \frac{1}{2} * n * \widehat{\boldsymbol{\beta}}^T\boldsymbol{\Omega^{-1}}\widehat{\boldsymbol{\beta}} + \sum_{i=1}^{P}[\gamma_i ln\pi(\alpha_i) + (1-\gamma_i)ln(1-\pi(\alpha_i)] \\
&= C - \frac{1}{2}ln|\boldsymbol{\Omega}| + \frac{m}{2}ln(\tau_\beta) + \frac{n}{2}\widehat{\boldsymbol{\beta}}^T\boldsymbol{\Omega^{-1}}\widehat{\boldsymbol{\beta}} + \sum_{i=1}^{P}[\gamma_i ln\pi(\alpha_i) + (1-\gamma_i)ln(1-\pi(\alpha_i)] \,.
\end{aligned}
\tag{12}
$$

The above log conditional posterior likelihood of $\gamma$ will be used in the Metropolis-Hastings (MCMC) algorithm [8] to make Bayesian inference for $\gamma$.

##### 1.5 GWAS summary data

Although the BFGWAS_QUANT model is developed for assuming the availability of individual-level GWAS data, the above log conditional posterior likelihood of $\gamma$ (12) can be calculated using reference LD $\boldsymbol{R}$, GWAS summary data $\widehat{\boldsymbol{\beta}}$, and GWAS sample size $n$. As shown in (9) and (10), $\boldsymbol{\Omega} = \boldsymbol{R} + \tau_\beta \boldsymbol{I_{m\times m}}$ can be obtained with a reference genotype correlation matrix $\boldsymbol{R}$, and the marginal effect sizes $\widehat{\boldsymbol{\beta}}$ can be obtained from single variant test Z-score statistics.

#### 2 EM-MCMC Algorithm

The EM-MCMC algorithm are implemented in BFGWAS_QUANT as follows:

(i) Fix $\tau_\beta$ as a value within $(0, 1]$, e.g., $1$ for using GWAS summary data. Fix $\alpha_0$ as a value within $[-13.8, -9]$, e.g., $-13.8$ to assume a sparse genetic causal architecture. Set initial values for enrichment coefficients $\{\alpha_j = 0, j = 1, ..., J\}$ that are shared by all genetic variants;

(ii) Generate $\boldsymbol{\pi}$ based on the hierarchical logistical model with most recent $\boldsymbol{\alpha}$ values and annotation data $\boldsymbol{A}$ as in (2),

$$\pi_i = \frac{e^{\boldsymbol{A_i'\alpha}}}{1 + e^{\boldsymbol{A_i'\alpha}}},\ i = 1, \cdots, p.$$

(iii) E-step: Conditioning on the most recent values of $\boldsymbol{\pi}$, estimate variant-specific parameters $(\boldsymbol{\beta}, E[\boldsymbol{\gamma}])$ by implementing MCMC per genome-block;

(iv) M-step: Conditioning on the estimates of $(\boldsymbol{\beta}, E[\boldsymbol{\gamma}])$ from the previous E-step, update $\{\alpha_j = 0, j = 1, ..., J\}$ by their MAPs, maximizing the expected log-posterior-likelihood functions as described in Section 2.2 [7];

(v) Repeat the EM steps (ii), (iii) and (iv) 3-5 times until the MAPs of $\{\alpha_j = 0, j = 1, ..., J\}$ converge. Estimates of $(\boldsymbol{\beta}, E[\boldsymbol{\gamma}])$ from the last E-step will be taken as the final estimates of genetic effect sizes and corresponding causal posterior probabilities (CPPs).

Hyper parameter values for $(\tau_\beta, \alpha_0)$ could be tuned by running the EM-MCMC algorithm with multiple choices of values for $(\tau_\beta, \alpha_0)$. One could select optimal hyper parameters based on the metrics of the fine-mapped results (e.g., total number of significant variants with CPP $> 0.1$), the magnitudes of Bayesian effect size estimates (which should be of similar magnitude as the marginal effect sizes), and regression $R^2$.

##### 2.1 MCMC sampling scheme

The MCMC sampling strategy is implemented per genome-block $k$ for estimating $(\boldsymbol{\beta}_k, E[\boldsymbol{\gamma_k}])$, conditioning on shared hyper parameters $(\boldsymbol{\alpha}, \tau_\beta)$. Details of the EM-MCMC algorithm was described in the supplementary note of the BFGWAS_CAT paper [1].

##### 2.2 MAP for enrichment coefficients

Conditioning on the indicator variable $\boldsymbol{\gamma}$, the conditional posterior density function (i.e., posterior likelihood) of $\boldsymbol{\alpha} = (\alpha_0, \alpha_1, ..., \alpha_j)$ can be derived from the joint posterior distribution (4) as follows:

$$P(\boldsymbol{\alpha}|\boldsymbol{\gamma}, \boldsymbol{A}) \propto P(\boldsymbol{\gamma}|\boldsymbol{\alpha}, \boldsymbol{A})P(\boldsymbol{\alpha}) = \prod_{i=1}^{p} P(\gamma_i|\boldsymbol{\alpha}, \boldsymbol{A_i})P(\boldsymbol{\alpha}),$$

where $\gamma_i|(\boldsymbol{\alpha}, \boldsymbol{A_i}) \sim Bernoulli(\pi_i)$, with $\pi_i = \frac{e^{\boldsymbol{A_i}\boldsymbol{\alpha}}}{1+e^{\boldsymbol{A_i}\boldsymbol{\alpha}}}$ and $\boldsymbol{\alpha} \sim MVN(0, \boldsymbol{I_J})$.

That is,

$$P(\boldsymbol{\alpha}|\boldsymbol{\gamma}, \boldsymbol{A}) \propto \prod_{i=1}^{p} \left[ \left( \frac{e^{\boldsymbol{A}_i'\boldsymbol{\alpha}}}{1 + e^{\boldsymbol{A}_i'\boldsymbol{\alpha}}} \right)^{\gamma_i} \left( 1 - \frac{e^{\boldsymbol{A}_i'\boldsymbol{\alpha}}}{1 + e^{\boldsymbol{A}_i'\boldsymbol{\alpha}}} \right)^{1-\gamma_i} \right] \frac{1}{\sqrt{(2\pi)^J}} \exp\left( -\frac{\boldsymbol{\alpha}'\boldsymbol{\alpha}}{2} \right)$$

$$\propto \prod_{i=1}^{p} \left[ \left( \frac{e^{\boldsymbol{A}_i'\boldsymbol{\alpha}}}{1 + e^{\boldsymbol{A}_i'\boldsymbol{\alpha}}} \right)^{\gamma_i} \left( 1 - \frac{e^{\boldsymbol{A}_i'\boldsymbol{\alpha}}}{1 + e^{\boldsymbol{A}_i'\boldsymbol{\alpha}}} \right)^{1-\gamma_i} \right] \exp\left( -\frac{\boldsymbol{\alpha}'\boldsymbol{\alpha}}{2} \right). \quad (13)$$

The expected log-posterior-likelihood function of $\boldsymbol{\alpha}$ is as follow:

$$
\begin{aligned}
l(\boldsymbol{\alpha}) &= E_{\gamma}[ln(P(\boldsymbol{\alpha}|\boldsymbol{\gamma}, \boldsymbol{A}))] \\
&\propto \sum_{i=1}^{p} \left[\hat{\gamma}_i ln\left(\frac{e^{\boldsymbol{A}_i'\boldsymbol{\alpha}}}{1+e^{\boldsymbol{A}_i'\boldsymbol{\alpha}}}\right) + (1-\hat{\gamma}_i) ln\left(1 - \frac{e^{\boldsymbol{A}_i'\boldsymbol{\alpha}}}{1+e^{\boldsymbol{A}_i'\boldsymbol{\alpha}}}\right)\right] - \frac{\boldsymbol{\alpha}'\boldsymbol{\alpha}}{2} \\
&\propto \sum_{i=1}^{p} \left[\hat{\gamma}_i ln\left(\frac{e^{\boldsymbol{A}_i'\boldsymbol{\alpha}}}{1+e^{\boldsymbol{A}_i'\boldsymbol{\alpha}}}\right) + \hat{\gamma}_i ln\left(1+e^{\boldsymbol{A}_i'\boldsymbol{\alpha}}\right) + ln\left(\frac{1}{1+e^{\boldsymbol{A}_i'\boldsymbol{\alpha}}}\right)\right] - \frac{\boldsymbol{\alpha}'\boldsymbol{\alpha}}{2} \\
&\propto \sum_{i=1}^{p} \left[\hat{\gamma}_i ln\left(e^{\boldsymbol{A}_i'\boldsymbol{\alpha}}\right) - ln\left(1+e^{\boldsymbol{A}_i'\boldsymbol{\alpha}}\right)\right] - \frac{\boldsymbol{\alpha}'\boldsymbol{\alpha}}{2} \\
&\propto \sum_{i=1}^{p} \left[\hat{\gamma}_i \boldsymbol{A}_i'\boldsymbol{a} - ln(1+e^{\boldsymbol{A}_i'\boldsymbol{a}})\right] - \frac{\boldsymbol{\alpha}'\boldsymbol{\alpha}}{2} \qquad (14)
\end{aligned}
$$

Since an analytical solution no longer exist for maximizing the above expected log-posterior-likelihood function (14) with respect to $\boldsymbol{\alpha}$, we utilize the nonlinear optimization method Broyden-Fletcher-Goldfarb-Shanno (BFGS) algorithm [9] as implemented in the *optimx()* function in the R library "optimx" [10] to estimate $\boldsymbol{\alpha}$. The gradient vector and hessian matrix of $l(\boldsymbol{\alpha})$ are given by

$$
\begin{aligned}
\frac{dl(\boldsymbol{\alpha})}{d\boldsymbol{\alpha}} &= \sum_{i=1}^{p} \left[\hat{\gamma}_i \boldsymbol{A}_i' - \left(\frac{e^{\boldsymbol{A}_i'\boldsymbol{\alpha}}}{1+e^{\boldsymbol{A}_i'\boldsymbol{\alpha}}}\right)\boldsymbol{A}_i'\right] - \boldsymbol{\alpha}' \\
&= \sum_{i=1}^{p} \left[\hat{\gamma}_i \boldsymbol{A}_i' - \left(1+e^{-\boldsymbol{A}_i'\boldsymbol{\alpha}}\right)^{-1}\boldsymbol{A}_i'\right] - \boldsymbol{\alpha}' \quad ;
\end{aligned} \qquad (15)
$$

$$
\begin{aligned}
\frac{d^2 l(\boldsymbol{\alpha})}{d\boldsymbol{\alpha} d\boldsymbol{\alpha}'} &= -\sum_{i=1}^{p} \left[\frac{e^{-\boldsymbol{A}_i'\boldsymbol{\alpha}}}{(1+e^{-\boldsymbol{A}_i'\boldsymbol{\alpha}})^2}(\boldsymbol{A}_i\boldsymbol{A}_i')\right] - \boldsymbol{I} \\
&= -\sum_{i=1}^{p} \left[\frac{e^{\boldsymbol{A}_i'\boldsymbol{\alpha}}}{2e^{\boldsymbol{A}_i'\boldsymbol{\alpha}} + e^{2\boldsymbol{A}_i'\boldsymbol{\alpha}} + 1}(\boldsymbol{A}_i\boldsymbol{A}_i')\right] - \boldsymbol{I} \quad .
\end{aligned} \qquad (16)
$$

#### 2.3 Fisher information of enrichment coefficients

Fisher information of $\boldsymbol{\alpha}$ can be derived from the second derivatives of the respective expected log-posterior-likelihood functions (16) as follows:

$$
\text{FisherInfo}(\boldsymbol{\alpha}) = -E_{\alpha}\left[\frac{e^{\boldsymbol{A}_i'\boldsymbol{\alpha}}}{2e^{\boldsymbol{A}_i'\boldsymbol{\alpha}} + e^{2\boldsymbol{A}_i'\boldsymbol{\alpha}} + 1}(\boldsymbol{A}_i\boldsymbol{A}_i')\right] + \boldsymbol{I}. \qquad (17)
$$

Here, $\alpha$ can be taken as values of their last estimates obtained in the last M step to obtain estimated Fisher information, which can be used to construct $95\%$ confidence intervals of enrichment coefficients. By the asymptotic-normality of MAP, as n $\rightarrow \infty$, the distribution of a MAP estimate converges to a multivariate normal distribution with mean equals to the true parameter value and covariance matrix equals to the inverse of the corresponding Fisher information.

#### 3 Software

Within the BFGWAS_QUANT software, C++ programming scripts are used to generate an executable file for the E-step (MCMC algorithm) and an R script is used for the M-step, which are wrapped together by a Makefile. Makefile script is generated by a Perl script and enables parallel computation through job management. This software is freely available at GitHub (`https://github.com/yanglab-emory/BFGWAS_QUANT`).
